# Adaptive hub reorganization distinguishes cognitive preservation from decline in epilepsy

**DOI:** 10.64898/2026.08.17.26360463

**Authors:** Tamjid Imtiaz, Alfredo Lucas, Emily Zhang, Mariam Josyula, Nina Petillo, Daniel J. Zhou, Mallory Mckee, Joel M. Stein, Kathy A. Lawler, Sandhitsu Das, Kathryn A. Davis

## Abstract

Cognitive impairment affects up to 80% of patients with drug resistant epilepsy (DRE), yet the basis for this impairment in patients with otherwise comparable disease characteristics remains poorly understood. Prior work has largely focused on identifying focal nodes responsible for cognitive decline, leaving the broader network reorganization associated with cognitive preservation poorly characterized. In this study, we hypothesized that the brain’s capacity to reorganize its functional network hubs, rather than the degree of underlying pathology, distinguishes cognitively resilient from cognitively impaired patients.

We studied a retrospective cohort of 105 DRE patients and 60 healthy controls who underwent resting-state functional neuroimaging. DRE patients were stratified into epilepsy cognitively neutral (ECN) and epilepsy cognitively impaired (ECI) subgroups based on comprehensive neuropsychological profiling spanning both domain-general and domain-specific levels. The subgroups did not differ in key disease characteristics including epilepsy duration, age of onset, seizure lateralization, and lesion status (*p*>0.05). We characterized hub organization across the whole brain, canonical functional networks and subcortical levels and summarized each subject’s functional reorganization using the hub disruption index. We found that whole brain topology is preserved in both groups whereas disruption concentrates in the salience network and dissociates within subcortical structures with reduced hippocampal node strength in both groups and increased thalamic node strength, with the latter more pronounced with cognitive burden. Inter-network connectivity shifted from focal, selective up-regulation in ECN to diffuse hyperconnectivity in ECI. Critically, the hub disruption index (HDI) for centrality separated the groups where the ECN group showed the greatest redistribution of centrality from canonical hubs towards alternative relay regions whereas ECI demonstrated comparatively little reorganization (ECN vs ECI: *d=*0.52, *p*=0.029; Bonferroni corrected). The same pattern held within individual domains, with greater hub reorganization in patients whose language and memory function was preserved.

These cross-sectional findings link cognitive impairment in epilepsy to a reduced capacity for adaptive hub reorganization rather than to pathology alone. Because the HDI for centrality is computable at the individual level, it may offer an objective imaging biomarker to complement neuropsychological testing, aid identification of patients at risk for cognitive decline, and inform prognostic counseling and surgical planning in DRE.

## Introduction

Cognitive impairment is one of the most prevalent comorbidities of epilepsy, affecting up to 80% of patients with drug resistant epilepsy in at least one neuropsychological domain and often severely impacting their quality of life^1,2^. Deficits can span multiple domains of cognitive function and are rarely confined to the structures classically linked to seizure generation^3^. Cognition in epilepsy is heterogeneous as it varies among patients with comparable disease characteristics, with the extent of distributed network abnormality scaling with the degree of cognitive compromise^4,5^. A recent effort has been made to provide an empirically grounded taxonomy for resolving this heterogeneity into well-defined cognitive phenotypes^6,7^. This phenotyping highlights a clinically important question: when the underlying epilepsy is comparable, what allows one brain to preserve cognition while another cannot, and does the answer lie in how its network reorganizes?

Recent evidence from clinical, electrophysiological, and neuroimaging studies has established epilepsy as a disorder of distributed brain networks rather than of an isolated focus^8–10^. Resting state functional magnetic resonance imaging (rs-fMRI) combined with graph theoretical analysis provides a quantitative framework to define these complex large scale networks and describes how efficiently information is integrated into the connectome and segregated into smaller modules^8^. Within this framework, the topology of the connectome has shown promising performance to track cognitive dysfunction in previous studies in TLE,^4,11^ thereby providing the methodological foundation of this study in characterizing and distinguishing the epilepsy phenotypes.

Highly-connected, interlinked, topologically central regions known as ‘hubs’ play a critical role in characterizing the network property by mediating the long-distance communication that underpins integrative connectivity and adaptive behavior^12–14^. In epilepsy, graph theoretical studies have repeatedly demonstrated the reorganization of hub architecture including disruption of canonical hubs and emergence of new hubs in formerly peripheral regions^15–17^. These changes have been observed in canonical resting state functional networks^18–20^ as well as in subcortical networks^21–23^. Importantly, hub-level reorganization is not always visible in measures of overall connectivity strength; rather, a network can preserve its aggregate functional connectivity while substantially redistributing where its hubs reside, so that connectivity-based and topology-based descriptions of the same network may diverge.

A growing body of work indicates that preserved cognition in the presence of epileptogenic pathology is not a passive state but an actively maintained one which recruits alternative regions and reconfigures the supportive network circuitry^24–26^. These studies resonate with the concept of the brain’s capacity to sustain function by flexibly reorganizing network resources against accumulating insult^27^. Under this view, topological reorganization may be a signature of successful compensation rather than of damage alone, and cognitive impairment may reflect, at least in part, a failure to reconfigure. Capturing this coordinated reorganization at the individual level requires a summary measure sensitive to the loss of control at the canonical hubs and gain of control in the peripheral regions. The hub disruption index, which was introduced to quantify the radical reorganization of functional hubs, has the capability to capture how far an individual’s nodal topology departs from the healthy pattern^28^. It has been applied across disorders of consciousness, stroke and lateralized focal epilepsy^16,28,29^; however, to our knowledge, it has not been used to ask whether hub reorganization distinguishes cognitive phenotypes within epilepsy.

Here, we address this gap by asking whether the brain’s capacity to reorganize its network hubs distinguishes preserved and impaired cognition in epilepsy. Specifically, we ask: does the capacity for hub reorganization determine whether cognition is preserved or impaired? Stratifying patients with drug-resistant focal epilepsy by cognitive phenotype into a cognitively neutral and cognitively impaired group and comparing both to healthy controls, we use rs-fMRI and a graph-theoretical approach to summarise each patient’s reorganization and answer this question. We show that the capacity for hub reorganization separates cognitively neutral from impaired patients in epilepsy such that impairment reflects a failure to reconfigure rather than the underlying pathology only. We further show that this adaptive reorganization relates not only to overall cognitive resilience but also to cognitive preservation of individual domains. Together, these findings identify reorganized hub topology, rather than preserved connectivity per se, as a network signature of cognitive resilience in epilepsy.

## Materials and methods

### Participants

We retrospectively studied 105 drug-resistant epilepsy patients (51 females, 54 males; mean age: 36.0 ± 12.3 years) from the Epilepsy Center at the University of Pennsylvania, all of whom were evaluated for surgical candidacy. Written informed consent was obtained from all participants prior to participation, and the study was approved by the institutional review board at the University of Pennsylvania (IRB#819126). Seventy four patients had temporal and 11 patients had frontal lobe epilepsy. Fifty six patients had left sided and 21 had right sided lateralization. Seizure localization and lateralization information was determined by the multidisciplinary participants in the Penn Epilepsy Surgical Conference incorporating available multimodal data (e.g., semiology, brain MRI, FDG-PET, MEG, scalp EEG, intracranial EEG). We also included 60 healthy controls (HC, 29 Female, 31 Male; mean age: 30.4 ± 9.4) in this study. Individual patient-specific information is provided in **Supplementary Table 1**.

### Cognition assessment

All the patients in the epilepsy cohort underwent neuropsychological test batteries as part of their clinical care spanning four cognitive domains including language, memory, executive function, and visual spatial perception. Of 105 epilepsy subjects, 82 completed assessments across all four domains and were included in the domain general cognitive classification analysis. All of the neuropsychological test scores were standardized to demographically corrected normative data and converted to t-scores^30^. Domain assignment of each of the tests followed published guidelines of International Classification of Cognitive Disorders in Epilepsy (IC-CoDE)^6^. Cognitive impairment within a given test was defined as t ≤ 35 as described in the guidelines. A domain was considered impaired when two or more constituent measures met this threshold. Given the statistical constraints of the dataset, participants were classified into two broad phenotypes: epilepsy cognitively neutral (all four domains intact) and epilepsy cognitively impaired (impairment in one or more domains), rather than the four IC-CoDE phenotypes. The patients were further analyzed at a specific domain level. Information including Neuropsychological measures for each domain is provided in **Supplementary Table 2**.

### Image Acquisition

Three different fMRI acquisition protocols were followed (fMRI protocols A, B and C), due to institutional changes in scanner and acquisition standards throughout the duration of this study. fMRI images for protocol A were acquired using a Siemens 3T Magnetom Trio scanner with the following parameters: 3.0 mm isotropic voxel size, echo time (TE)/repetition time (TR) = 30/500 ms, multiband factor of 6, and a 7 min acquisition time. fMRI images for protocol B were acquired using a Siemens 3T Magnetom Trio scanner with the following parameters: 2 mm isotropic voxel size, TE/TR = 37/800 ms, multiband factor of 8, and 6 min acquisition time. Finally, fMRI images for protocol C were acquired using a Siemens 3T Magnetom PrismaFit scanner with the following parameters: 2 mm isotropic voxel size, TE/TR = 37/800 ms, multiband factor of 8, and 9 min acquisition time. For all the protocols, T1 weighted images were acquired with a sagittal, 208-slice MPRAGE sequence, TE/TR = 2.24/2400 ms, inversion time (TI) =1060 ms, field-of-view (FOV) = 256mm, with a 0.8 mm isotropic voxel size. The distribution of subjects across the protocols is provided in **Supplementary Table 1**.

### Functional image processing

We used fMRIPrep^31^ to perform brain extraction and segmentation of individual T1-weighted (T1w) images, registration of rsfMRI volumes to individual T1w and 152 non-linear asymmetric 2009 Montreal Neurological Institute (MNI) version c (MNI152NLin2009cAsym) template space, and time-series confound estimation. Then, we used the fMRIPrep output data as our input to the xcpEngine post-processing pipeline^32^ for 36-parameter confound regression, demeaning, detrending, and temporal filtering. Complete details of the pre-processing pipeline used in this study are included in **Supplementary Methods**.

### Functional connectivity

After the preprocessing step, we measured the average BOLD time series for each voxel within each parcel of the Schaefer 200 atlas (200 cortical and 56 subcortical parcels)^33^. We created a 256 × 256 functional connectivity matrix by computing Pearson correlations between mean time series at each parcel. To correct for technical variability and potential batch effects introduced by the three different acquisition protocols, we harmonized the subject-level functional connectivity (FC) matrices using NeuroCombat^34^. We included the group assignments (HC and Epilepsy) as a covariate to preserve in NeuroCombat. To assess the effect of harmonization, data distributions before and after correction are depicted in a two-dimensional principal component space (Supplementary Figure **S1**).

### Network metrics

We measured network topological properties at three levels: whole-brain, inter-network, and intra-network. At the whole-brain level, analyses were performed across proportional thresholds of network density ranging from 10% to 50% in 2% increments. We established a density range of 10–50% to mitigate network fragmentation at the lower bound and exclude confounding random associations at the upper bound^35^. We assessed four graph-theoretical metrics: node strength, betweenness centrality, clustering coefficient, and efficiency^36^. Node strength reflects the overall connectivity of a node by summing the weights of all its connections. Betweenness centrality quantifies the extent to which a node lies on the shortest paths between other nodes, indicating its role in network communication. The clustering coefficient measures the interconnectedness of a node’s neighbors and reflects local network segregation. Efficiency characterizes the capacity for information transfer within the network, with higher values indicating more efficient communication^37^. In the inter and intra-network analysis, we chose Yeo’s seven functional canonical networks: default mode network (DMN), frontoparietal network (FPN), salience attention network (SAN), dorsal Attention Network (DAN), limbic network (LN), visual network (VN), and somatosensory motor network (SMN) to measure their internal characteristics as well as their interaction with other networks^38^. We calculated the same network measures across proportional thresholds of network density ranging from 10% to 50% in 5% increments. We assessed the network measures across a range of connection densities to confirm that the results were robust and reproducible. For inter-network analysis, we calculated the average connectivity between network pairs across the same range of network densities used for the intra-network analysis. Additionally, we also measured the node strength in hippocampus and thalamus which are known subcortical regions involved in epilepsy and cognitive functions^39^. Nodal metrics were averaged across all nodes in the brain for the whole brain level analysis and across networks for intranetwork analysis, resulting in a single global average value for each metric across densities.

### Graph topological hub disruption indices

Hub disruption index calculation was carried out to identify any reorganization of the central nodes or hubs of a complex network compared to the healthy controls. We estimated the hub disruption index, *K*, for the graph metrics: disruption index for node strength (*K*_S_), betweenness centrality (*K*_BC_), clustering coefficient (*K*_CC_), and efficiency (*K_G_*_E_) by following methods described in Termenon *et al.*^28^. This measure allows us to summarize the abnormal profile of nodal connectivity and topological metrics of an individual subject in relation to the normative topology of the HC group. For each subject, we first subtracted the HC group mean graph metric from the same metric of the corresponding node in a given individual; next, we plotted this individual difference against the HC group mean. The hub disruption index, *K*, is then defined as the slope of a straight line fitted to the scatter plot following the linear regression (*y* = *Kx* + *b*), where *y* = subject - mean of HC; *x* = mean of HC; *b* = residual or intercept of the regression. Here, a negative K value indicates that canonical hub nodes lose connectivity while peripheral nodes abnormally gain it, reflecting a fundamental reorganization of network topology. Whereas, a positive K value indicates that hub locations are abnormally hyperconnected relative to the healthy brain, with canonical organization being reinforced. After computing the individual disruption indices, significant differences between groups in *K*_S_, *K*_BC_, *K*_CC_, and *K*_E_ were calculated at 30% network density. The rationale for choosing this threshold is that at 10% network density, networks tend to become disconnected at low densities, and the upper bound of 50%, since network properties at higher densities tend to randomness due to the inclusion of potentially confounding associations. Additionally, to verify if the network density has no impact on the hub disruption index, we calculated the same hub disruption indices across proportional thresholds of network density ranging from 10% to 50% in 5% increments. This analysis was conducted both in the overall cognition as well as specific domain level.

### Statistical Analysis

Group demographic and clinical characteristics were compared across HC, ECN and ECI groups. Continuous variables such as age, age at onset, epilepsy duration are reported as mean ± standard deviation and compared across the groups using the Kruskal-Wallis test. Binary variables such as sex, MRI lesional status, hippocampal sclerosis, and other lesional etiology are reported as count and percentage and were compared using a chi-squared test. Statistical significance was set at p<0.05. The clinical and demographic characteristics of the patients with domain general cognitive phenotypes are summarized in Table **1**. Only the ‘age’ variable reached significance here and was corrected in the subsequent analysis. To evaluate the significance of group level differences, a non parametric cluster permutation test was applied between group pairs (HC vs ECN, HC vs ECI, and ECN vs ECI) and contiguous suprathreshold clusters were identified^40^. A null distribution of maximum cluster sizes was generated from 5000 random permutations of group labels, and the permutation p value was identified as the proportion of permuted maximum cluster sizes exceeding the observed value, Statistical Significance was set at p<0.05. This analysis was conducted at three level: (1) whole brain, averaging nodal metrics across all 256 parcels; (2) network level, averaging metrics within each of the seven canonical Yeo networks across nine density steps (0.10 to 0.50 with 0.05 increment); and (3) subcortical, averaging metrics within the hippocampus and thalamus across the same density range as (2). Inter network functional connectivity was computed as the mean functional connectivity between each of the unique network pairs after proportional thresholding, across nine density steps (0.10 to 0.50 with 0.05 increment). For comparison across a fixed network density, the statistical significance was assessed using an independent sample t test and Bonferroni correction was used to correct for multiple comparisons. We considered Cohen’s *d* values above 0.2 to represent a small effect size, 0.5 to represent a moderate effect size, and above 0.8 to represent a large effect size^41^.

**Table 1.** Demographic and Clinical Information.

| Characteristic | Controls<br>(N=60) | ECN<br>(N=30) | ECI<br>(N=52) | p-value |
| --- | --- | --- | --- | --- |
| Age at scan, years | 30.4 ± 9.4 | 39.8 ± 14.3 | 35.3 ± 11.3 | 0.002 |
| Male sex (%) | 31 (51.7) | 11 (36.7%) | 29 (55.8%) | 0.2355 |
| Disease duration, years | — | 21.5 ± 16.3 | 15.6 ± 13.6 | 0.092 |
| Age at onset, years | — | 18.4 ± 13.4 | 19.7 ± 12.0 | 0.541 |
| Seizure lateralization |  |  |  | 0.810 |
| Left | — | 16 (55%) | 26 (50%) |  |
| Right | — | 8 (28%) | 18 (35%) |  |
| Other (Bilateral and inconclusive) | — | 5 (17%) | 8 (15%) |  |
| Seizure localization |  |  |  | 0.885 |
| Temporal | — | 22 (73%) | 36(69%) |  |
| Frontal | — | 3 (10%) | 5 (10%) |  |
| Other | — | 5 (17%) | 11 (21%) |  |
| MRI lesional, n (%) | — | 10 (34%) | 22 (42%) | 0.570 |
| MRI lesion type |  |  |  |  |
| MTS | — | 5 (17%) | 6 (12%) | 0.749 |
| Other | — | 5 (17%) | 16 (31%) | 0.252 |

**Abbreviations:**
ECI: Epilepsy Cognitively Impaired,
ECN: Epilepsy Cognitively Neutral,
MTS: Mesial Temporal Sclerosis

For hub disruption index, group differences in *K* at the single density level (0.3) were evaluated with independent sample t test and Bonferroni correction. To confirm these effects were not tied to an arbitrary density threshold, we computed all four HDIs across densities 0.10–0.50 as well with the same non parametric cluster permutation test as described before. Stabilities across densities were assessed using the same cluster permutation test described previously. Similar statistical analyses were conducted in the domain specific level as well. Associations with clinical variables were examined using non-parametric tests appropriate to variable type. Spearman rank correlations were used for continuous variables. Mann-Whitney U tests (two-sided) were used for binary variables. Kruskal-Wallis tests were used for categorical variables. Epilepsy-specific variables (all except age and sex) were tested only within the epilepsy cohort. All raw p-values across the full set of valid tests were corrected for multiple comparisons using the false discovery rate (FDR) procedure. Associations with FDR-corrected *p* < 0.05 were considered statistically significant.

## Results

### Global network topology properties are unaltered in ECN and ECI

Whole-brain functional network topology was broadly preserved across groups (ECN and ECI), with no statistically significant differences compared to HC observed at any connection density (10–50%) for node strength, clustering coefficient, global efficiency, or betweenness centrality (all p_perm_ > 0.07; Figure **S2**). Average node strength, global efficiency, and clustering scaled upward with increasing density, while betweenness centrality declined across all groups. Despite the lack of significance among the groups, some trends are observed in the network topology. Node strength was elevated in ECI throughout densities compared to HC and ECN (Figure **S2A**), while betweenness centrality was lowest in ECN, implying reduced hub-like integration in cognitively neutral epilepsy patients (Figure **S2B**). Clustering exhibited a density-dependent divergence, with ECN showing lower local segregation at lower densities but increased clustering at higher densities compared to ECI (Figure **S2C**).

### Altered salience attention network topology characterizes cognitive function in epilepsy from controls

Across the seven canonical resting-state networks, intra-network disruptions were more pronounced and widespread within the Salience Attention Network (SAN) in all four graph metrics. Node strength was significantly reduced in both ECN (*d* = 0.56, *p_bonf_* = 0.04) and ECI (*d* = 0.60, *p_bonf_* < 0.01) relative to HC (**Figure 2A**). Clustering coefficient was also significantly diminished in both ECN (*d* = 0.623, *p_bonf_* = 0.02) and ECI (*d* = 0.61, *p_bonf_* < 0.01) compared to HC (**Figure 2A**). Global efficiency followed a similar pattern, with significant reductions in ECN (d = 0.562, *p_bonf_*= 0.04) and ECI (d = 0.618. *p_bonf_* < 0.01) relative to HC, underscoring deficits in rapid network-wide communication (**Figure 2A**). Betweenness centrality followed the opposite trend, with elevations in both ECI (*d* = 0.5, *p_bonf_* = 0.03) and ECN (*d =* 0.6, *p_bonf_*= 0.03) compared to HC, reflecting increased hub-like routing capacity within the SAN (**Figure 2A**). Together, these findings suggest that the SAN is particularly significant in epilepsy-related cognitive function, with impairments emerging even in cognitively neutral subjects and becoming more severe with cognitive impairment. We found this to be significant even when analyzing the graph metrics across different network densities **(Figure S3).** Within the limbic network (LN), betweenness centrality was found higher in ECN (*d* = 0.63, *p_bonf_* < 0.05) compared to HC, indicating increased information flow within the LN specifically in cognitively neutral epilepsy subjects (**Figure 2A**). However, across network densities the effect was significant in both ECN and ECI groups. No significant differences were found among the two epilepsy cognitive phenotypes in the intra-network characteristics of the remaining five networks (Figure S3). Furthermore, given the critical role of subcortical structures in large-scale network regulation, we examined the node strength in the thalamus and hippocampus structures across groups. Hippocampal node strength was significantly reduced in both ECN (*d* = 0.81, *p_bonf_* < 0.01) and ECI (*d* = 1.1, *p_bonf_* < 0.001) compared to HC, indicating that hippocampal connectivity disruption is an early and sustained consequence of epilepsy, present even before overt cognitive impairment manifests and becoming more severe with cognitive decline. Thalamic node strength showed the opposite pattern, with elevations in ECN and further pronounced increments in ECI (*p_bonf_* < 0.001) relative to HC, suggesting a compensatory or pathological hyperconnectivity of the thalamus in the context of epilepsy (**Figure 2C**). Across different network densities in hippocampus and thalamus, significant node strength differences consistent with the single density level were found **(Figure S4)**. The thalamus, as a critical relay hub, appears particularly susceptible to epilepsy-related dysconnectivity, and its graded increase in node strength across cognitive groups may reflect progressive thalamocortical recruitment in response to widespread cortical dysfunction. Together, these subcortical findings reveal that while hippocampal node strength degrades with increasing cognitive burden, thalamic node strength increases, reinforcing the notion that disruption of subcortical-cortical circuits underlies epilepsy-related cognitive impairment through distinct and opposing mechanisms^42^.

**Figure 1.**
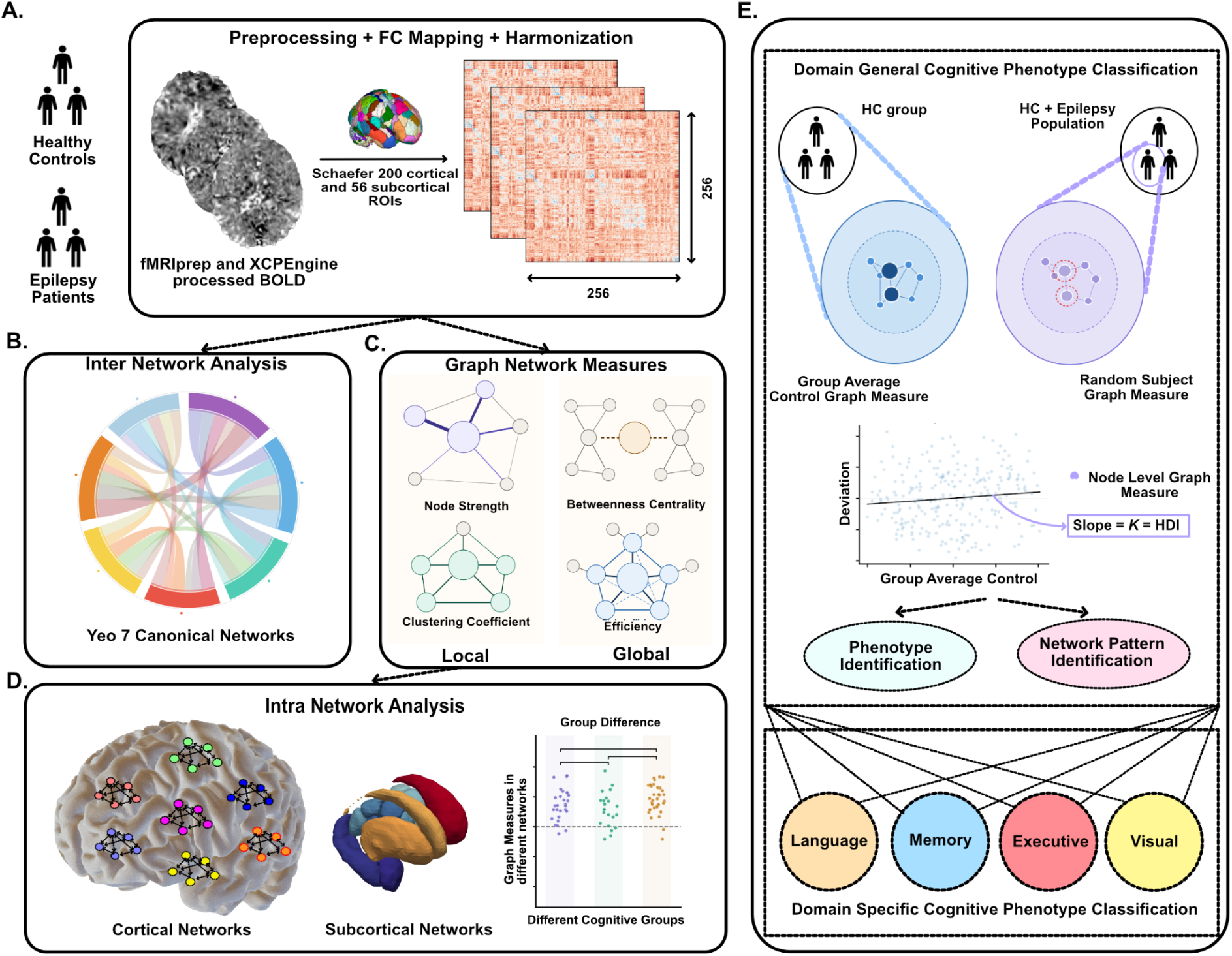
Methodological Overview. **(A)** BOLD signals were preprocessed, functional connectivity matrices were derived from the resulting time series, and data were harmonized across acquisition sites. (**B)** Average functional connectivity across Yeo’s seven canonical resting-state networks was compared across cognitive groups. (**C&D)** Graph-theoretical metrics were computed across cortical networks and subcortical regions and compared across cognitive phenotypes. (**E)** The hub disruption index (*K*) was derived by regressing individual subject graph metric values against the group-averaged healthy control profile. Controls show minimal deviation from this reference, whereas patients deviate systematically which is captured by the slope *K*. Group differences in *K* were compared across all graph metrics to characterize hub reorganization across epilepsy cognitive phenotypes, and this framework was subsequently extended to four cognitive domains. *FC = Functional Connectivity, HC = Healthy Controls, HDI = Hub Disruption Index.

**Figure 2.**
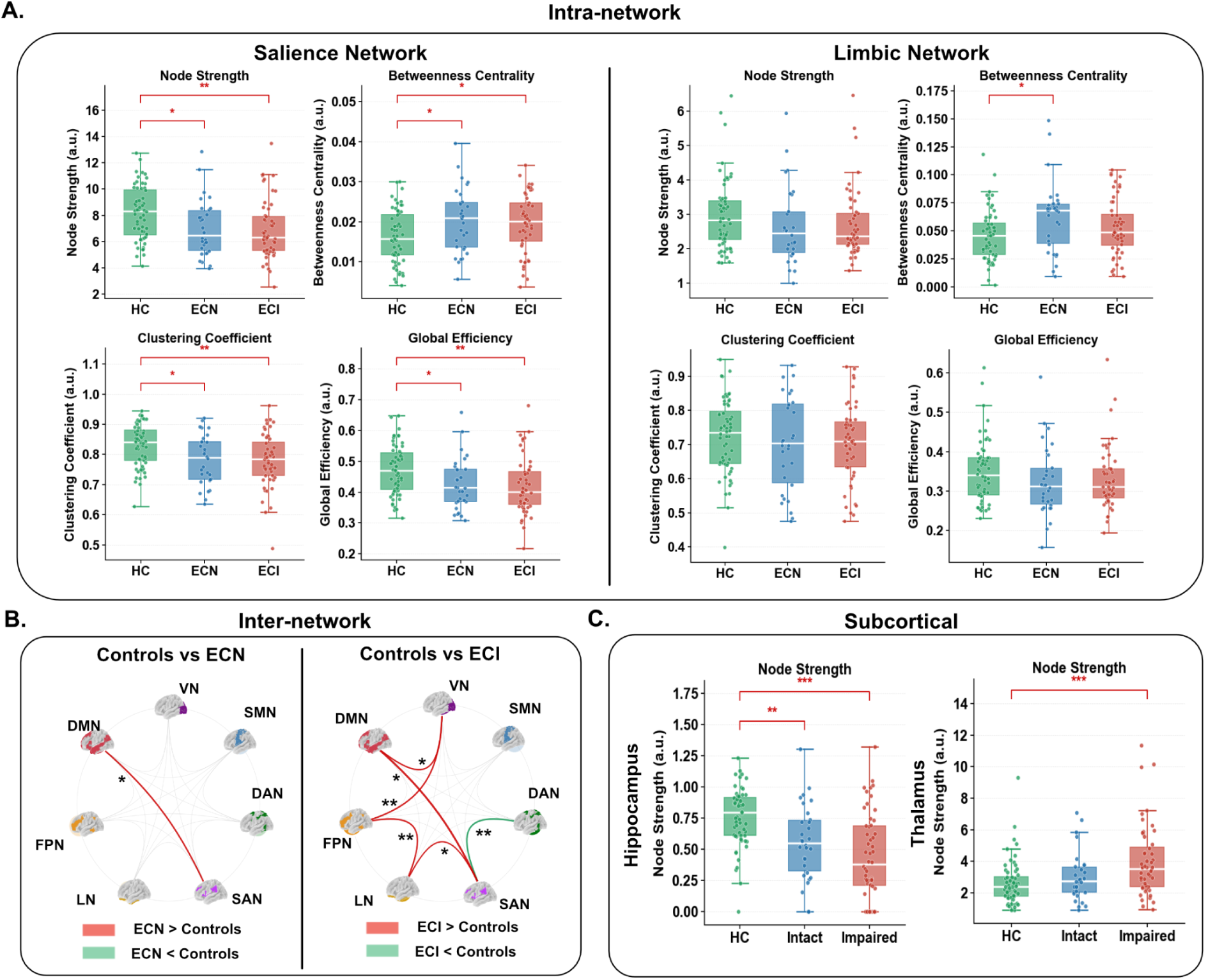
Graph-theoretic and node-level connectivity alterations across cognitive phenotypes in epilepsy. **(A)** Intra-network graph metrics for the salience (left) and limbic (right) networks, shown for HC (green), ECN (blue), and ECI (red). For each network, node strength, betweenness centrality, clustering coefficient, and global efficiency are plotted. (**B)** Inter-network connectivity differences relative to controls, shown as chord diagrams over the seven Yeo canonical functional networks. Red edges indicate connections stronger in patients than controls and green edges connections weaker than controls. (**C)** Subcortical node strength for the hippocampus (left) and thalamus (right) in HC versus cognitively neutral and impaired patients. DMN, default mode; VN, visual; SMN, somatomotor; DAN, dorsal attention; SAN, salience; LN, limbic; FPN, frontoparietal. \**p* < 0.05, \*\**p* < 0.01, \*\*\**p* < 0.001.

### Elevated inter-network connectivity in epilepsy reflects a compensatory-to-pathological shift in cognition

In the HC-versus-ECN comparison, the inter-network connection between default mode (DMN) and visual (VN) networks was significantly stronger (*p_bonf_* < 0.05) at lower network density (0.1), and the default mode-salience (DMN–SAN) connection was significantly elevated in ECN relative to HC (*p_bonf_*< 0.05) across a broad density range(0.15–0.5). This selective strengthening of connections between networks that do not typically interact was consistent with an early, focal compensatory up-regulation of inter-network connectivity. The ECI group showed a markedly different profile: relative to controls, ECI exhibited widespread increases in inter-network connectivity, most prominently among the DMN, frontoparietal (FPN), VN and SAN alongside reduced connectivity in the salience-dorsal attention (SAN–DAN) connection **(Figure S5, Figure 2B**). As network density increased, progressively more networks contributed to the increased-connectivity pattern, whereas the SAN–DAN decrease remained consistent across all densities. This hyperconnectivity among networks that normally retain relative functional independence may reflect a loss of network segregation rather than adaptive reorganization. Taken together, the inter-network findings suggest a mechanistic transition from focal compensatory hyperconnectivity in ECN to diffuse pathological hyperconnectivity in ECI, positioning inter-network integration as a sensitive marker of cognitive trajectories in epilepsy.

### Hub disruption in centrality can differentiate epilepsy related cognitive phenotypes

To compute the hub disruption index, we first derived node-level graph metrics for every subject and z-scored them across nodes before averaging within the reference control group (**Figure 3A**). For each individual, we then regressed every region’s deviation from this control mean against the control-group nodal profile. The resulting slope of that fit defines the HDI, a single value capturing how far a subject’s hub organization departs from the normative pattern for each of the four graph metrics (**Figure 3B**). HDI showed no association with age, age of onset, disease duration, lesional status, seizure laterality, sex, or etiology, indicating that it indexes network topology rather than clinical confounds **(Supplementary Table 2)**. Among the metrics, betweenness-centrality disruption (*K_BC_*) separated all three groups: HC vs ECI (d=0.50, *p_bonf_*<0.05), HC vs ECN (d=0.99, *p_bonf_*<0.001), and ECN vs ECI (*d*=0.52, *p_bonf_*<0.05), whereas *K_S_*, *K_CC_*, and *K_E_* distinguished patients from controls but not ECN from ECI (**Figure 3C**). Critically, ECN showed the most negative *K_BC_*, indicating the emergence of alternate relay hubs to reroute information and supporting our hypothesis that hub reorganization contributes to cognitive resilience. ECI, by contrast, stayed closer to the control slope and reorganized little suggesting that cognitive impairment reflects not pathology alone but a failure to topologically reconfigure, leaving the network unable to compensate.

**Figure 3:**
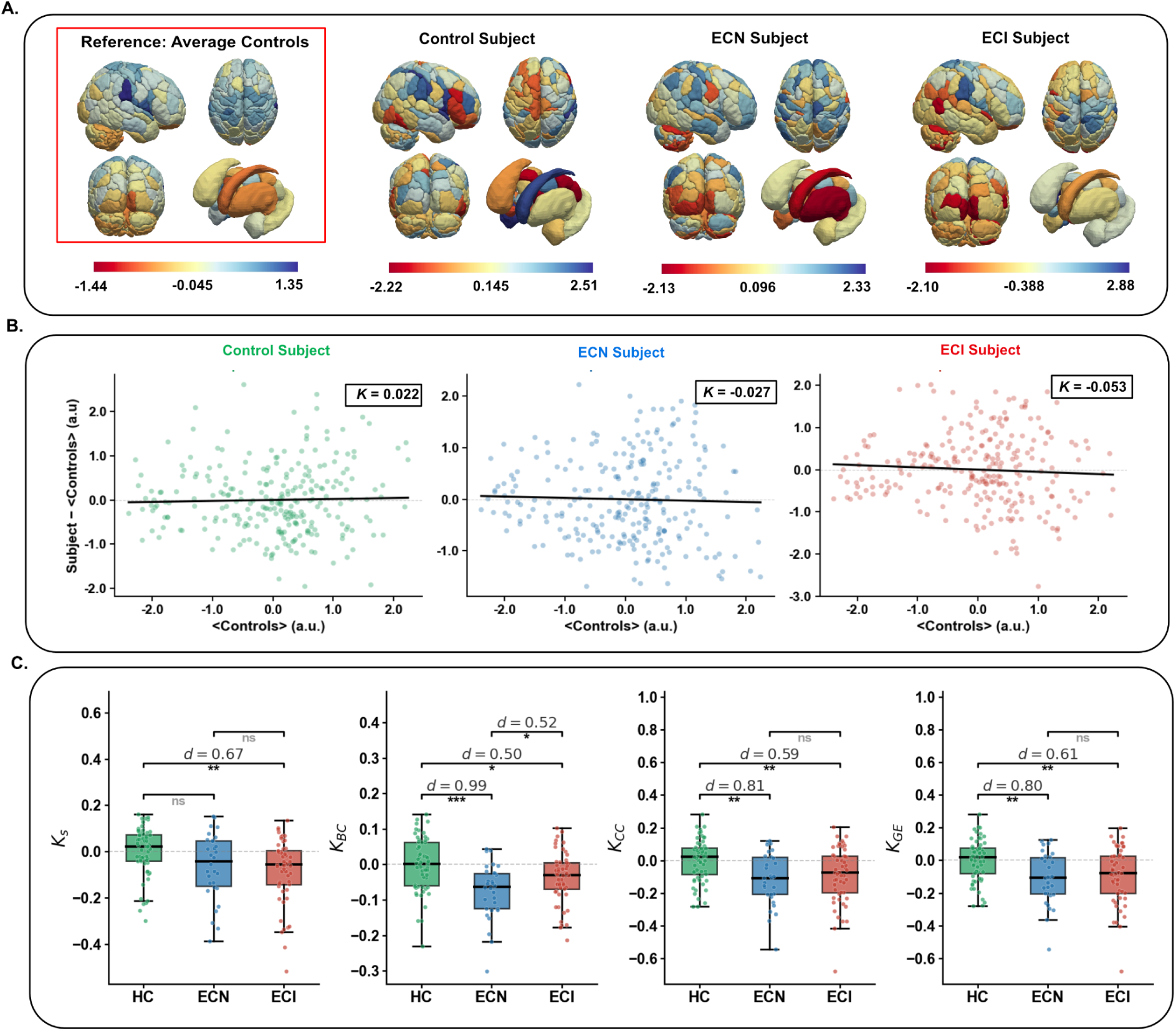
Hub disruption indices reveal graded topological reorganization across cognitive phenotypes. **(A)** Whole-brain maps of regional node strength (z-scored, arbitrary units) projected onto cortical and subcortical surfaces. The boxed reference map (left, red outline) shows the average control profile; the three adjacent maps show representative single subjects (a control, an ECN, and an ECI patient). **(B)** Hub disruption index for the three representative subjects in (A). For each region, the individual deviation from the control mean (subject − ⟨controls⟩) is plotted against the corresponding control mean (⟨controls⟩). The slope of the linear fit (black line) defines the hub disruption index (*K*) for the corresponding graph metric. **©** Group distributions of *K* derived from four graph metrics — node strength (*K*_S_), betweenness centrality (*K*_BC_), clustering coefficient (*K*_CC_), and global efficiency (*K*_GE_), for HC (green), ECN (blue), and ECI (red). Patient groups showed predominantly negative κ across metrics. Betweenness centrality showed the strongest separation of ECN from controls, exceeding the HC–ECI difference, consistent with pronounced topological reorganization in cognitively normal patients. Significance: **ns**, not significant; **\**p* < 0.05, \*\**p* < 0.01, \*\*\**p* < 0.001.**

### Spatial redistribution of betweenness centrality underlies hub disruption across epilepsy cognitive phenotypes

To localize the hub disruption in terms of betweenness centrality summarized by *K_BC_*, we decomposed each pairwise contrast into node-level contributions, plotting every region’s reference-group BC against its between-group difference (**Figure 4A–B, left)**. This consistently revealed two opposing node populations: high-BC regions in the reference group (red triangle) and low-BC regions that gained centrality (blue triangle). The simultaneous demotion of canonical hubs and promotion of formerly peripheral nodes is precisely what generates the negative *K_BC_*slope. The brain maps and bar plots representing the number of deviated nodes in the cortical and subcortical networks (**Figure 4, middle and right)** show where this redistribution occurs. In HC vs ECN (**Figure 4A**), the higher-BC nodes were concentrated in the somatomotor (SMN) and default-mode (DMN) networks, whereas the lower-centrality fell mainly on dorsal-attention (DAN) and frontoparietal (FPN) regions. In HC vs ECI (**Figure 4B**), the lower centrality pattern is dominated by the SMN and extended across visual (VN), DAN, LN, and FPN, while gains are centered on the SAN and DMN networks. Together, these maps suggest that the cognitively preserved group concentrates compensatory centrality gains in a sensorimotor–default-mode set, whereas the impaired group fails to sustain this targeted reallocation and disperses centrality across networks. To confirm these effects were not tied to an arbitrary density threshold, we recomputed all four HDIs across densities 0.10–0.50 (**Figure S6**). Although the hub disruption index in centrality was not overall significant across the whole range, it is still significant in the range (0.3-0.35). The significance difference observed in this middle range likely reflects a genuine effect rather than an artifact of threshold choice. It is because betweenness centrality depends on shortest path structure, making it sensitive to network density where too few edges destabilize the path and too many introduce redundant paths that weakens hub signals

**Figure 4.**
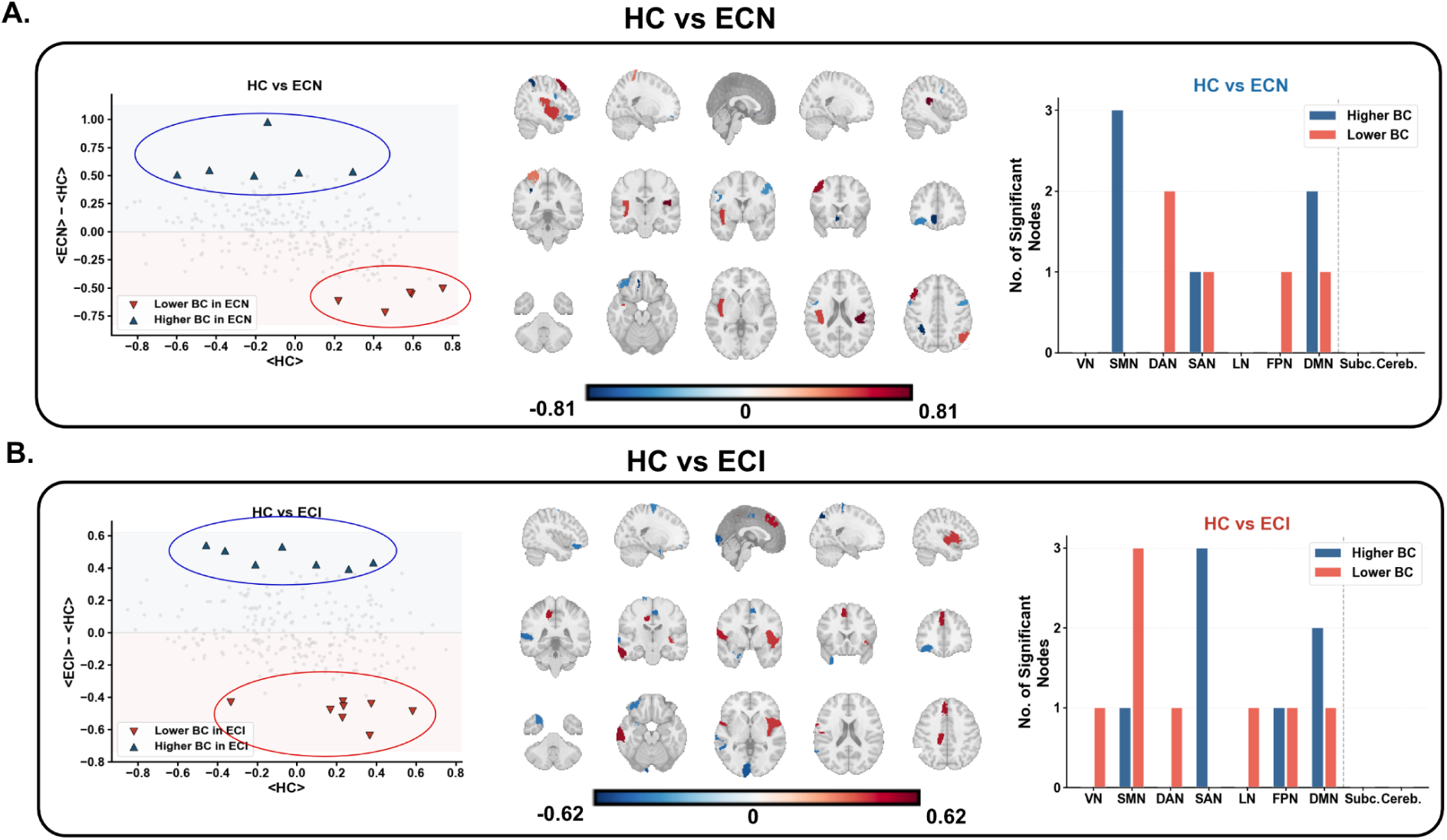
Node-level betweenness-centrality (BC) reorganization across pairwise group comparisons. **(A)** HC vs. ECN and (**B)** HC vs. ECI. For each comparison, the scatter (left) plots the regional betweenness-centrality (BC) difference between groups (y-axis: ⟨group 2⟩ − ⟨group 1⟩) against the reference group’s mean BC (x-axis), with each point a parcel. Gray points are non-significant; colored triangles mark parcels surviving statistical thresholding. Two opposing populations emerge: high-BC reference regions that lose centrality in the second group (downward triangles, lower-right) and low-BC reference regions that gain centrality (upward triangles, upper-left). The middle part of each panel projects the significant parcels onto sagittal, coronal, and axial MNI slices, colored by the signed BC difference (blue, higher in the second group; red, lower; per-comparison scales shown below). The bar charts (right) summarize the number of significant nodes per network — visual (VN), somatomotor (SMN), dorsal attention (DAN), salience (SAN), limbic (LN), frontoparietal (FPN), default mode (DMN), subcortical (Subc.), and cerebellar (Cereb.), split by direction (Higher BC, blue; Lower BC, red).

### Betweenness-centrality hub disruption generalizes across cognitive domains

To investigate whether the betweenness-centrality hub-disruption signature observed at the broader level of cognition generalized to specific cognitive domains, we recomputed *K*_BC_ after stratifying the patient groups by language, memory, executive function and visual spatial attention status (**Figure 5 and Figure S7**). In language and memory domains, the cognitively preserved subgroup deviated most from controls: *K*_BC_ was significantly more negative in patients with neutral language function (Lang-Neutral) than HC (*d* = 0.85, *p_bonf_* < 0.0001) and in patients with neutral memory function (Mem-Neutral) than HC (*d* = 0.92, *p_bonf_*< 0.0001), with smaller but significant shifts in the impaired subgroups (HC vs Lang-Impaired, *d* = 0.48; HC vs Mem-Impaired, *d* = 0.43; both *p_bonf_* < 0.05). The neutral and impaired subgroups were also separable (Lang-Neutral vs Lang-Impaired, *d* = 0.42; Mem-Neutral vs Mem-Impaired, *d* = 0.55; both *p_bonf_* < 0.05). In both domains, the neutral subgroup showed the greater topological reorganization, the same ordering seen at the overall domain level.

**Figure 5.**
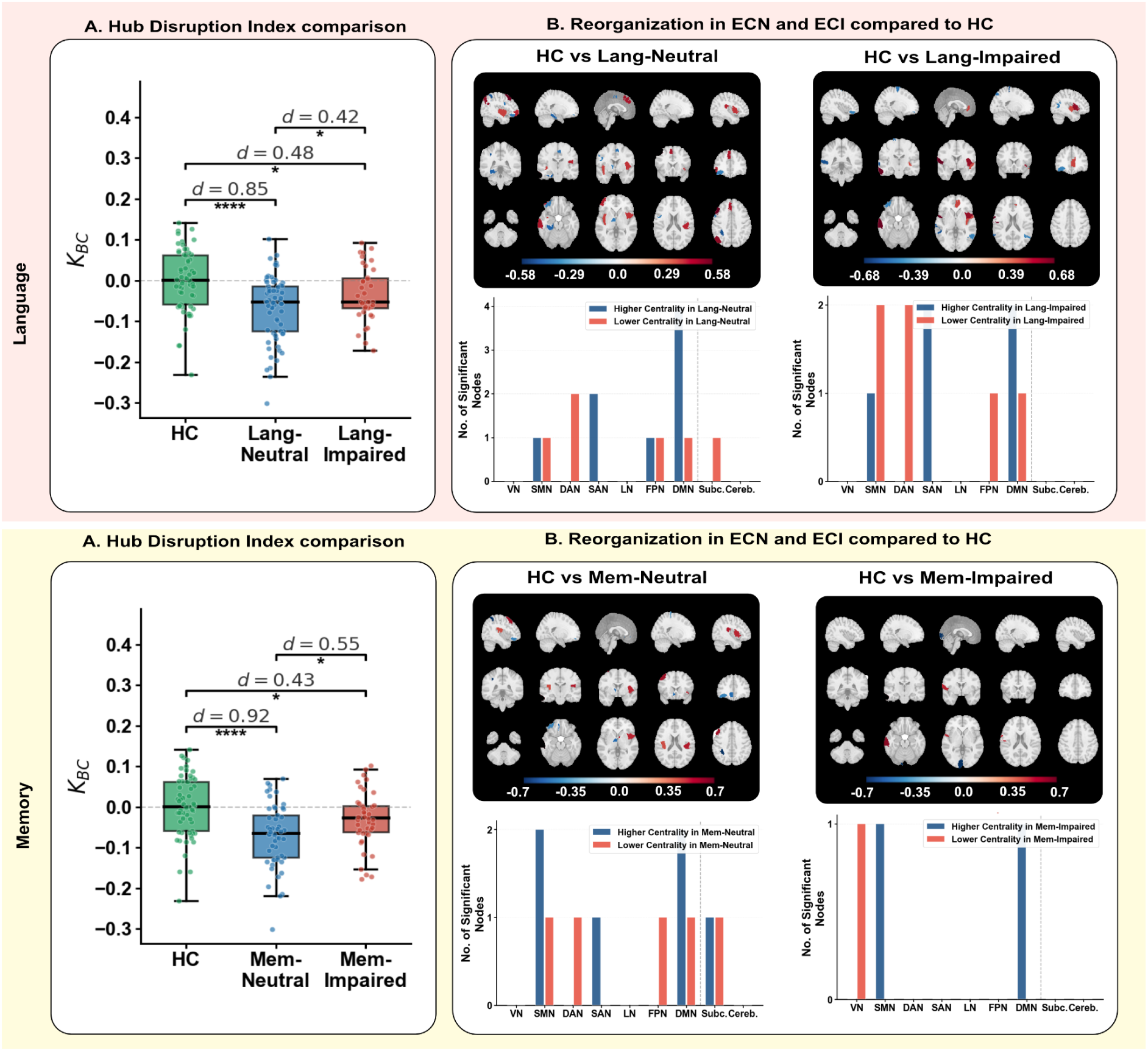
Domain-specific betweenness-centrality reorganization for language and memory phenotypes. Results are shown for two cognitive domains: language (top) and memory (bottom). In each domain, patients are split by domain-specific performance into an neutral and an impaired subgroup, each compared against the same healthy-control reference. (**A)** Group distributions of the betweenness-centrality hub disruption index (*K*_BC_) for HC, the neutral subgroup, and the impaired subgroup (language: Lang-Neutral *n* = 67, Lang-Impaired *n* = 38; memory: Mem-Neutral *n* = 53, Mem-Impaired *n* = 46). (**B)** Node-level reorganization relative to controls for each subgroup. Brain panels project parcels with significant betweenness-centrality differences onto sagittal, coronal, and axial MNI slices, colored by direction (blue, higher centrality in the patient subgroup; red, lower). The accompanying bar charts count significant nodes per network — visual (VN), somatomotor (SMN), dorsal attention (DAN), salience (SAN), limbic (LN), frontoparietal (FPN), default mode (DMN), subcortical (Subc.), and cerebellar (Cereb.) — split by direction. Significance: \**p* < 0.05, \*\*\*\**p* < 0.0001.

Node-level mapping (**Figure 6B and D**) localized these effects. In the language domain, the preserved subgroup showed gained-centrality nodes concentrated in the default-mode (DMN) and salience (SAN) networks, with losses falling mainly on the dorsal-attention (DAN) network. In the impaired subgroup, centrality was lost in the somatomotor (SMN) and DAN networks and gained in the SAN; overall, the impaired group showed greater SMN and DAN losses than the neutral subgroup. For memory, the preserved subgroup showed centrality losses in the DMN relative to controls, offset by gains in the SMN and SAN networks; the impaired subgroup exhibited similar SMN and DMN changes but markedly less reorganization overall. Together, these patterns support the idea that more extensive and widespread network reorganization underlies the preservation of cognitive function. We did the same hub disruption analysis in the other three graph metrics however, they are not significant across the three cognitive groups (**Figure S7**).

Finally, we performed the same analysis for executive function (EF) and visual-spatial perception (VSP) (**Figure S8 and S9**). In executive function, both subgroups were significantly disrupted relative to controls (EF-Neutral vs HC: *d*=0.71, *p*<0.001 and EF-Impaired: *d* = 1.03, *p*<0.01). Although the difference was not significant, a more negative betweenness-centrality hub disruption was observed in the impaired subgroup than in the neutral one, suggesting that executive dysfunction, unlike language or memory impairment, may be accompanied by pronounced rather than diminished hub reorganization. In neither domain were the neutral and impaired subgroups statistically separable, and node-level gains in the preserved subgroups again clustered in the SMN, SAN, and DMN. For VSP, significant hub disruption relative to controls was confined to the preserved subgroup across all four indices, whereas the impaired subgroup did not differ from controls on any metric.

## Discussion

In a retrospective cohort of 105 patients with drug resistant focal epilepsy and 60 healthy controls, stratified by neuropsychological phenotype into cognitively neutral (ECN) and cognitively impaired (ECI) subgroups, we characterized network reorganization across whole-brain, network, and nodal scales and summarized it at the level of the individual using the hub disruption index. Four key findings emerged. First, whole-brain network topology was statistically indistinguishable across groups, yet pronounced reorganization emerged at finer scales. Second, intra-network disruption was concentrated in the salience network and accompanied by an opposing subcortical pattern of reduced hippocampal and elevated thalamic node strength that scaled with cognitive impairment. Third, inter-network connectivity shifted from a focal, selective up-regulation in ECN to a diffuse, widespread hyperconnectivity in ECI. Fourth, and most centrally, the betweenness-centrality hub disruption index separated all three groups, with the ECN group showing the most negative values while the ECI group with comparatively minimal reorganization. This signature was independent of clinical confounds and recurred within the language and memory domains. Together, these results support a model in which cognitive resilience in epilepsy appears to be associated with an actively reconfigured network topology rather than a passively preserved one, and in which impairment reflects, in part, a failure to reconfigure^24,43^.

The salience network was the principal cortical network of disruption, showing reduced node strength, clustering and efficiency alongside elevated betweenness centrality in both patient groups. A network that is internally weakened and less segregated, yet carries disproportionate shortest-path traffic, has become a more critical relay bottleneck for inter-system communication. This interpretation is consistent with the salience network’s established role as the dynamic switch that toggles between the default-mode and executive systems^44–46^. The graded severity from ECN to ECI suggests that salience-network compromise begins before overt impairment and deepens with it, consistent with reports linking salience-network dysfunction to attentional and broader cognitive deficits in epilepsy^20^. The subcortical findings add a complementary mechanism where hippocampal node strength fell while thalamic node strength rose with cognitive burden. The opposing direction of these two subcortical effects argues against a single global dysconnectivity gradient and instead implicates circuit-specific mechanisms underlying epilepsy-related cognitive change.

The inter-network results trace the same compensatory-to-pathological trajectory at the level of system interactions. Prior functional studies have consistently reported reduced connectivity, significantly lateralizing to seizure onset zones and most pronounced to limbic and default systems^19^. Against this backdrop, the inter-network hyperconnectivity we observed stands out. It likely reflects a different, system-level process than the focal disconnection near the seizure focus that dominates the existing literature: local decreases at the focus mark the primary pathology, while the distributed increases we observe may reflect secondary reorganization. Consistent with this in ECN, hyperconnectivity was selective and largely confined to default-mode coupling with the salience and visual systems, consistent with targeted, focal up-regulation. In ECI, hyperconnectivity became widespread, recruiting default-mode, frontoparietal, visual and salience systems, while the salience–dorsal-attention link weakened. This reconfiguration is naturally framed by the segregation–integration balance that healthy cognition requires. Optimal performance depends on the brain’s ability to maintain and flexibly switch between segregated and integrated configurations, and both excessive integration and excessive segregation are associated with poorer cognition^47–49^. In this view, the selective integration seen in ECN may preserve that balance, whereas the diffuse, indiscriminate hyperconnectivity of ECI represents a collapse of modular boundaries and integration without specificity that is maladaptive rather than compensatory.

The hub reorganization signature reappeared when patients were re-stratified by single-domain status, with the language and memory-preserved subgroups again deviating most from controls. This domain-level reproduction strengthens the claim that targeted hub redistribution underpins preserved functionality. Importantly, the pattern was not uniform across domains. In the executive-function analysis, the ordering trended in the opposite direction, with the impaired subgroup showing the more reorganized rather than the preserved hub organization, and in the visuospatial analysis, only the preserved subgroup differed from controls. We interpret this heterogeneity cautiously, as these contrasts did not reach significance for the subgroup separation, but it suggests that the compensation-by-redistribution mechanism is most pertinent to language and memory, the domains most tightly bound to the temporal-lobe networks that dominate this cohort.

Since hub disruption index was unrelated to age, age at onset, disease duration, lesional status, seizure laterality, sex or etiology, it appears to index network topology rather than clinical confounds, which is the property required of a candidate biomarker. An externally validated single, subject-level metric that distinguishes cognitively neutral from impaired patients and that may reflect latent compensatory capacity rather than current performance alone, could complement labour-intensive neuropsychological batteries and, with validation, inform the prognostication of cognitive trajectories, including the risk of post-operative decline in patients evaluated for surgery^50,51^.

We have several limitations in our study. First, the design is cross-sectional in a drug resistant focal epilepsy cohort and, thus, we cannot establish that hub reorganization is causally protective rather than an epiphenomenon of preserved cognition. Longitudinal and, ideally, interventional data are needed to test directionality. Second, statistical constraints obliged us to collapse the IC-CoDE taxonomy into a binary intact/impaired contrast, which discarded phenotype granularity. Finally, we had limited statistical power to characterize several cognitive domains: executive function and visual spatial perception.

## Conclusion

Our study presents a novel insight into the functional reorganization associated with cognitive preservation in epilepsy. We demonstrated that cognitively preserved patients show the most reorganization, redistributing betweenness centrality away from the usual hubs and toward alternative relay regions, while impaired patients have comparatively less reorganized networks. These findings recast cognitive resilience in epilepsy as an actively reconfigured network rather than a passively preserved one, and cognitive impairment as a failure to reconfigure rather than the result of pathology alone. Future work would include longitudinal studies where we can establish whether hub reorganization precedes and protects against cognitive decline or simply accompanies it, and ultimately whether it can forecast how cognition changes over the course of the disease including epilepsy surgical interventions. Furthermore, combining functional connectivity with structural connectivity may also begin to explain why some brains reorganize while others cannot. Our findings provide preliminary evidence that an objective, patient level marker of reorganizational capacity could complement neuropsychological testing and help guide cognitive prognosis and surgical planning in drug resistant epilepsy.

## Data Availability

All data produced in the present study are available upon reasonable request to the Principal Investigator.

## Data availability

All code and dependencies used for this manuscript are available at https://github.com/tamjidimtiaz/Adaptive-Cognitive-Reorganization-in-Epilepsy. Data are available upon reasonable request from Principal Investigator Kathryn A. Davis.

## Acknowledgements

We thank all the participants for their participation in this study. We also thank all other members and staff of the Center for Neuroengineering and Therapeutics for their continued help and support in this work.

