## Supplementary Material for "Adaptive hub reorganization distinguishes cognitive preservation from decline in epilepsy"

### **Neuroimaging Preprocessing**

Preprocessing was performed using fMRIPrep 20.2.3^1^, which is based on Nipype 1.6.1^2^.

#### **Anatomical data preprocessing**

For each subject, T1w images were corrected for intensity non-uniformity (INU) with N4BiasFieldCorrection^3^, distributed with ANTs 2.3.3^4^. The T1w-reference was then skull-stripped with a Nipype implementation of the antsBrainExtraction.sh workflow (from ANTs), using OASIS30ANTs as target template. Brain tissue segmentation of cerebrospinal fluid (CSF), whitematter (WM) and gray-matter (GM) was performed on the brain-extracted T1w using FAST (FSL 5.0.9)^5^. A T1w-reference map was computed after registration of 2 T1w images (after INUcorrection) using mri_robust_template (FreeSurfer 6.0.1)^6^. Brain surfaces were reconstructed using FreeSurfer’s recon-all, and the brain mask estimated previously was refined with a custom variation of the method to reconcile ANTs-derived and FreeSurfer-derived segmentations of the cortical gray-matter of Mindboggle^7^. Volume-based spatial normalization to one standard space (MNI152NLin2009cAsym) was performed through nonlinear registration with antsRegistration (ANTs 2.3.3), using brain-extracted versions of both T1w reference and the T1w template. The following template was selected for spatial normalization: ICBM 152 Nonlinear Asymmetrical template version 2009c^8^.

#### **Functional data preprocessing**

For the single BOLD run of each subject, the following preprocessing was performed. First, a reference volume and its skull-stripped version were generated using a custom methodology of fMRIPrep. A B0-nonuniformity map (or fieldmap) was estimated based on a phase-difference map calculated with a dual-echo GRE (gradient-recall echo) sequence, processed with a custom workflow of SDCFlows inspired by the epidewarp.fsl script and further improvements in HCP Pipelines^9^. The fieldmap was then co-registered to the target EPI (echo-planar imaging) reference run and converted to a displacements field map (amenable to registration tools such as ANTs) with FSL’s fugue and other SDCflows tools. Based on the estimated susceptibility distortion, a corrected EPI (echo-planar imaging) reference was calculated for a more accurate co-registration with the anatomical reference. The BOLD reference was then co-registered to the T1w reference using bbregister (FreeSurfer) which implements boundary-based registration^10^. Co-registration was configured with six degrees of freedom. Head-motion parameters with respect to the BOLD reference (transformation matrices, and six corresponding rotation and translation parameters) are estimated before any spatiotemporal filtering using mcflirt (FSL 5.0.9)^11^. BOLD runs were slicetime corrected using 3dTshift from AFNI 20160207^12^. The BOLD time-series (including slicetiming correction when applied) were resampled onto their original, native space by applying a single, composite transform to correct for head-motion and susceptibility distortions. These resampled BOLD time-series will be referred to as preprocessed BOLD in original space, or just preprocessed BOLD. The BOLD time-series were resampled into standard space, generating a preprocessed BOLD run in MNI152NLin2009cAsym space. First, a reference volume and its skull-stripped version were generated using a custom methodology of fMRIPrep. Several confounding time-series were calculated based on the preprocessed BOLD: framewise displacement (FD), DVARS and three region-wise global signals. FD was computed using Power’s formulation^13^ (absolute sum of relative motions) and Jenkinson’s approach (relative root mean square displacement between affines)^11^. FD and DVARS are calculated for each functional run, both using their implementations in Nipype^2^. The three global signals are extracted within the CSF, the WM, and the whole-brain masks. Additionally, a set of physiological regressors were extracted to allow for component-based noise correction (CompCor)^14^. Principal components are estimated after high-pass filtering the preprocessed BOLD time-series (using a discrete cosine filter with 128s cut-off) for the two CompCor variants: temporal (tCompCor) and anatomical (aCompCor). tCompCor components are then calculated from the top 2% variable voxels within the brain mask. For aCompCor, three probabilistic masks (CSF, WM and combined CSF+WM) are generated in anatomical space. The implementation differs from that of Behzadi et al.^14^ in that instead of eroding the masks by 2 pixels on BOLD space, the aCompCor masks are subtracted from a mask of pixels that likely contain a volume fraction of GM. This mask is obtained by dilating a GM mask extracted from the FreeSurfer’s aseg segmentation, and it ensures components are not extracted from voxels containing a minimal fraction of GM. Finally, these masks are resampled into BOLD space and binarized by thresholding at 0.99 (as in the original implementation). Components are also calculated separately within the WM and CSF masks. For each CompCor decomposition, the k components with the largest singular values are retained, such that the retained components’ time series are sufficient to explain 50 percent of variance across the nuisance mask (CSF, WM, combined, or temporal). The remaining components are dropped from consideration. The head-motion estimates calculated in the correction step were also placed within the corresponding confounds file. The confound time series derived from head motion estimates and global signals were expanded with the inclusion of temporal derivatives and quadratic terms for each. Frames that exceeded a threshold of 0.5 mm FD or 1.5 standardised DVARS were annotated as motion outliers. All resamplings can be performed with a single interpolation step by composing all the pertinent transformations (i.e. head-motion transform matrices, susceptibility distortion correction when available, and co-registrations to anatomical and output spaces). Gridded (volumetric) resamplings were performed using antsApplyTransforms (ANTs), configured with Lanczos interpolation to minimize the smoothing effects of other kernels. Non-gridded (surface) resamplings were performed using mri_vol2surf (FreeSurfer).

The output of the fMRIPrep pre-processing pipeline was subsequently input into the xcpEngine post-processing pipeline^15^. The xcpEngine post-processing pipeline is a self-contained software that allows the rapid and reproducible implementation of tools necessary for calculating functional connectivity, while also allowing for benchmarking pipeline performance using a wide array of benchmarking pipelines^16^. xcpEngine is built to use the output of the fMRIPrep pipeline as an input, therefore, many of the pipeline steps use metrics explicitly calculated by fMRIPrep. Briefly, the steps implemented in the xcpEngine pipeline for each subject, were as follows. First, regressors for artifactual signals were calculated from the 4D time series of each subject using the confound2 module. The regressed parameters calculated by this module included motion realignment parameters (3 rotational and 3 translational) necessary for realigning each volume in the time series to a reference volume; the mean white matter and cerebrospinal fluid time series over all voxels^17^ , with tissue segmentations determined by the fMRIPrep; the mean time series signal across the whole brain^13^; the temporal derivative of motion parameters, which encodes the relative displacement of the brain from one volume of the timeseries to the next^17^; and finally, the second power of each of the previously mentioned regressors was also included, to account for potential noise that is proportional to higher powers of motion and nuisance regressors (a total of 36 regression parameters). After estimating the regressors, demeaning and detrending, followed by temporal filtering was carried out in both the BOLD timeseries and the regressors, using the regress module. The timeseries and regressors were detrended using a 2nd order polynomial. A first order forward-backward bandpass Butterworth filter, with passband 0.01-0.10Hz was implemented, allowing both high frequency noise, and very-low-frequency drift to be eliminated. The filtered regressors were fitted to the filtered BOLD timeseries data using multiple linear regression. Any variance in the BOLD timeseries explained by the regressors was discarded from the timeseries, whereas the unexplained variance was left as the final filtered timeseries.

**Supplementary Table 1: Subject Demographics**

| **Sub ID** | **Subject Type** | **Sex** | **Age at scan (Range)** | **Protocol** | **Seizure Localization** | **Seizure Lateralization** | **MTS (1=present, 0=Non-MTS)** | **MRI Lesion** | **Age at onset (Range)** |
| --- | --- | --- | --- | --- | --- | --- | --- | --- | --- |
| 1 | Epilepsy | Female | 61-70 | A | Temporal | L | 1 | Y | 1-10 |
| 2 | Epilepsy | Female | 11-20 | A | Temporal | R | 1 | Y | 11-20 |
| 3 | Epilepsy | Female | 11-20 | A | Multi-focal | R | 0 | Y | 11-20 |
| 4 | Epilepsy | Male | 51-60 | B | Temporal | R | 0 | N | 21-30 |
| 5 | Epilepsy | Female | 31-40 | A | Temporal | L | 0 | N | 1-10 |
| 6 | Epilepsy | Female | 11-20 | B | Frontal | L | 0 | N | 11-20 |
| 7 | Epilepsy | Female | 41-50 | A | Temporal | L | 0 | N | 21-30 |
| 8 | Epilepsy | Female | 21-30 | A | Non-localizable | Inconclusive | 0 | Y | 1-10 |
| 9 | Epilepsy | Male | 31-40 | B | Frontal | R | 0 | N | 1-10 |
| 10 | Epilepsy | Female | 51-60 | B | Temporal | L | 0 | N | 11-20 |
| 11 | Epilepsy | Female | 51-60 | B | Temporal | R | 0 | N | 1-10 |
| 12 | Epilepsy | Male | 31-40 | B | Temporal | L | 0 | N | 21-30 |
| 13 | Epilepsy | Male | 31-40 | B | Non-localizable | Inconclusive | 0 | N | 1-10 |
| 14 | Epilepsy | Male | 41-50 | B | Temporal | Bilateral | 0 | N | 31-40 |
| 15 | Epilepsy | Female | 41-50 | B | Non-localizable | L | 0 | N | 21-30 |
| 16 | Epilepsy | Female | 41-50 | C | Temporal | Bilateral | 0 | N | 11-20 |
| 17 | Epilepsy | Male | 41-50 | C | Temporal | R | 1 | Y | 11-20 |
| 18 | Epilepsy | Male | 21-30 | C | Temporal | L | 0 | N | 11-20 |
| 19 | Epilepsy | Female | 31-40 | C | Temporal | R | 0 | Y | 21-30 |
| 20 | Epilepsy | Male | 31-40 | C | Temporal | L | 0 | N | 1-10 |
| 21 | Epilepsy | Female | 21-30 | C | Non-localizable | Inconclusive | 0 | N | 11-20 |
| 22 | Epilepsy | Female | 31-40 | C | Temporal | L | 1 | Y | 21-30 |
| 23 | Epilepsy | Female | 41-50 | C | Temporal | L | 1 | Y | 31-40 |
| 24 | Epilepsy | Female | 21-30 | C | Temporal | L | 0 | N | 1-10 |
| 25 | Epilepsy | Male | 21-30 | C | Frontal | L | 0 | Y | 11-20 |
| 26 | Epilepsy | Female | 31-40 | C | Temporal | L | 0 | N | - |
| 27 | Epilepsy | Male | 21-30 | C | Frontal | L | 0 | N | 11-20 |
| 28 | Epilepsy | Male | 21-30 | C | Non-localizable | Inconclusive | 0 | N | 11-20 |
| 29 | Epilepsy | Female | 21-30 | C | Frontal | L | 0 | N | 1-10 |
| 30 | Epilepsy | Male | 31-40 | C | Non-localizable | L | 0 | N | 21-30 |
| 31 | Epilepsy | Female | 61-70 | C | Temporal | L | 0 | Y | 51-60 |
| 32 | Epilepsy | Female | 21-30 | C | Temporal | L | 0 | Y | 11-20 |
| 33 | Epilepsy | Male | 21-30 | C | Temporal | L | 0 | N | 11-20 |
| 34 | Epilepsy | Male | 31-40 | C | Temporal | L | 0 | Y | 21-30 |
| 35 | Epilepsy | Male | 21-30 | C | Temporal | L | 0 | Y | 21-30 |
| 36 | Epilepsy | Male | 21-30 | C | Temporal | R | 0 | N | 11-20 |
| 37 | Epilepsy | Female | 51-60 | C | Non-localizable | Inconclusive | 0 | N | 1-10 |
| 38 | Epilepsy | Female | 21-30 | C | Temporal | L | 0 | Y | 21-30 |
| 39 | Epilepsy | Female | 21-30 | C | Temporal | L | 0 | N | 11-20 |
| 40 | Epilepsy | Female | 31-40 | C | Temporal | L | 0 | N | 1-10 |
| 41 | Epilepsy | Male | 41-50 | C | Non-localizable | Inconclusive | 0 | N | 11-20 |
| 42 | Epilepsy | Male | 31-40 | C | Multi-focal | L | 0 | Y | 21-30 |
| 43 | Epilepsy | Female | 41-50 | C | Temporal | R | 0 | N | 1-10 |
| 44 | Epilepsy | Female | 41-50 | C | Temporal | L | 1 | Y | 1-10 |
| 45 | Epilepsy | Female | 11-20 | C | Central | L | 0 | N | 11-20 |
| 46 | Epilepsy | Female | 41-50 | C | Temporal | L | 0 | N | 1-10 |
| 47 | Epilepsy | Male | 21-30 | C | Multi-focal | L | 0 | Y | 21-30 |
| 48 | Epilepsy | Male | 61-70 | C | Temporal | R | 0 | Y | 21-30 |
| 49 | Epilepsy | Male | 31-40 | C | Temporal | Bilateral | 0 | Y | 21-30 |
| 50 | Epilepsy | Male | 21-30 | C | Temporal | L | 0 | Y | 21-30 |
| 51 | Epilepsy | Male | 31-40 | C | Temporal | L | 0 | N | 11-20 |
| 52 | Epilepsy | Female | 61-70 | C | Temporal | Bilateral | 0 | N | 31-40 |
| 53 | Epilepsy | Male | 11-20 | C | Temporal | L | 0 | Y | 1-10 |
| 54 | Epilepsy | Female | 41-50 | C | Temporal | L | 0 | N | 31-40 |
| 55 | Epilepsy | Female | 31-40 | C | Temporal | L | 1 | Y | 31-40 |
| 56 | Epilepsy | Male | 21-30 | C | Multi-focal | L | 0 | N | 11-20 |
| 57 | Epilepsy | Male | 31-40 | C | Temporal | Bilateral | 0 | N | 31-40 |
| 58 | Epilepsy | Male | 21-30 | C | Temporal | R | 0 | Y | 1-10 |
| 59 | Epilepsy | Male | 31-40 | C | Temporal | L | 0 | Y | 21-30 |
| 60 | Epilepsy | Female | 41-50 | C | Temporal | R | 1 | Y | 1-10 |
| 61 | Epilepsy | Male | 31-40 | C | Temporal | R | 1 | Y | 11-20 |
| 62 | Epilepsy | Female | 51-60 | C | Temporal | R | 0 | Y | 51-60 |
| 63 | Epilepsy | Male | 31-40 | C | Temporal | R | 0 | Y | 21-30 |
| 64 | Epilepsy | Female | 21-30 | C | Temporal | L | 1 | Y | 1-10 |
| 65 | Epilepsy | Male | 21-30 | C | Central | L | 0 | Y | 1-10 |
| 66 | Epilepsy | Female | 21-30 | C | Frontal | L | 0 | Y | 1-10 |
| 67 | Epilepsy | Female | 31-40 | C | Temporal | R | 0 | N | - |
| 68 | Epilepsy | Male | 41-50 | C | Temporal | R | 0 | Y | 31-40 |
| 69 | Epilepsy | Male | 21-30 | C | Temporal | L | 1 | Y | 1-10 |
| 70 | Epilepsy | Male | 31-40 | C | Frontal | R | 0 | N | 1-10 |
| 71 | Epilepsy | Male | 31-40 | C | Temporal | L | 1 | Y | 21-30 |
| 72 | Epilepsy | Male | 21-30 | C | Temporal | L | 0 | N | 11-20 |
| 73 | Epilepsy | Female | 41-50 | C | Multi-focal | Inconclusive | 1 | Y | 1-10 |
| 74 | Epilepsy | Male | 21-30 | C | Multi-focal | R | 0 | N | 1-10 |
| 75 | Epilepsy | Male | 41-50 | C | Temporal | L | 0 | N | 31-40 |
| 76 | Epilepsy | Female | 31-40 | C | Frontal | R | 0 | N | 11-20 |
| 77 | Epilepsy | Male | 11-20 | C | Temporal | L | 0 | N | 11-20 |
| 78 | Epilepsy | Male | 31-40 | C | Temporal | R | 0 | N | 21-30 |
| 79 | Epilepsy | Male | 21-30 | C | Frontal | L | 0 | Y | 11-20 |
| 80 | Epilepsy | Female | 41-50 | C | Temporal | R | 0 | Y | 31-40 |
| 81 | Epilepsy | Female | 41-50 | C | Temporal | L | 0 | N | 41-50 |
| 82 | Epilepsy | Female | 31-40 | C | Temporal | R | 0 | Y | 21-30 |
| 83 | Epilepsy | Female | 21-30 | C | Temporal | R | 0 | N | 11-20 |
| 84 | Epilepsy | Female | 21-30 | C | Temporal | L | 0 | N | 11-20 |
| 85 | Epilepsy | Female | 21-30 | C | Temporal | R | 0 | N | 1-10 |
| 86 | Epilepsy | Male | 21-30 | C | Temporal | R | 1 | Y | 11-20 |
| 87 | Epilepsy | Female | 21-30 | C | Temporal | R | 0 | N | 21-30 |
| 88 | Epilepsy | Female | 31-40 | C | Temporal | L | 0 | N | 21-30 |
| 89 | Epilepsy | Male | 61-70 | C | Temporal | L | 0 | N | 11-20 |
| 90 | Epilepsy | Male | 61-70 | C | Frontal | R | 0 | N | 1-10 |
| 91 | Epilepsy | Female | 21-30 | C | Occipital |  | 0 | N | 1-10 |
| 92 | Epilepsy | Female | 31-40 | C | Temporal | R | 0 | Y | 21-30 |
| 93 | Epilepsy | Male | 21-30 | C | Non-localizable | L | 0 | N | 1-10 |
| 94 | Epilepsy | Male | 41-50 | C | Temporal | R | 0 | N | 31-40 |
| 95 | Epilepsy | Female | 31-40 | C | Temporal | L | 0 | N | 21-30 |
| 96 | Epilepsy | Male | 21-30 | C | Frontal | R | 0 | Y | 1-10 |
| 97 | Epilepsy | Male | 41-50 | C | Non-localizable | Inconclusive | 0 | Y | 21-30 |
| 98 | Epilepsy | Male | 21-30 | C | Non-localizable | Inconclusive | 1 | Y | 21-30 |
| 99 | Epilepsy | Male | 31-40 | C | Temporal | L | 0 | N | 21-30 |
| 100 | Epilepsy | Male | 51-60 | C | Temporal | L | 1 | Y | 11-20 |
| 101 | Epilepsy | Male | 31-40 | C | Temporal | L | 1 | Y | 1-10 |
| 102 | Epilepsy | Male | 31-40 | C | Temporal | Bilateral | 1 | Y | 31-40 |
| 103 | Epilepsy | Male | 31-40 | C | Temporal | R | 0 | N | 11-20 |
| 104 | Epilepsy | Female | 21-30 | C | Temporal | L | 0 | Y | 11-20 |
| 105 | Epilepsy | Female | 51-60 | C | Temporal | Bilateral | 0 | N | 21-30 |
| 106 | Controls | Female | 21-30 | A | NA | NA | NA | NA | NA |
| 107 | Controls | Female | 11-20 | A | NA | NA | NA | NA | NA |
| 108 | Controls | Male | 21-30 | A | NA | NA | NA | NA | NA |
| 109 | Controls | Male | 21-30 | B | NA | NA | NA | NA | NA |
| 110 | Controls | Female | 31-40 | B | NA | NA | NA | NA | NA |
| 111 | Controls | Male | 31-40 | B | NA | NA | NA | NA | NA |
| 112 | Controls | Male | 21-30 | B | NA | NA | NA | NA | NA |
| 113 | Controls | Male | 31-40 | B | NA | NA | NA | NA | NA |
| 114 | Controls | Female | 31-40 | C | NA | NA | NA | NA | NA |
| 115 | Controls | Male | 21-30 | C | NA | NA | NA | NA | NA |
| 116 | Controls | Female | 21-30 | C | NA | NA | NA | NA | NA |
| 117 | Controls | Male | 61-70 | C | NA | NA | NA | NA | NA |
| 118 | Controls | Female | 31-40 | C | NA | NA | NA | NA | NA |
| 119 | Controls | Male | 21-30 | C | NA | NA | NA | NA | NA |
| 120 | Controls | Male | 21-30 | C | NA | NA | NA | NA | NA |
| 121 | Controls | Male | 21-30 | C | NA | NA | NA | NA | NA |
| 122 | Controls | Male | 31-40 | C | NA | NA | NA | NA | NA |
| 123 | Controls | Male | 21-30 | C | NA | NA | NA | NA | NA |
| 124 | Controls | Female | 31-40 | C | NA | NA | NA | NA | NA |
| 125 | Controls | Male | 21-30 | C | NA | NA | NA | NA | NA |
| 126 | Controls | Male | 21-30 | C | NA | NA | NA | NA | NA |
| 127 | Controls | Female | 21-30 | C | NA | NA | NA | NA | NA |
| 128 | Controls | Male | 21-30 | C | NA | NA | NA | NA | NA |
| 129 | Controls | Female | 21-30 | C | NA | NA | NA | NA | NA |
| 130 | Controls | Male | 11-20 | C | NA | NA | NA | NA | NA |
| 131 | Controls | Male | 21-30 | C | NA | NA | NA | NA | NA |
| 132 | Controls | Female | 21-30 | C | NA | NA | NA | NA | NA |
| 133 | Controls | Male | 21-30 | C | NA | NA | NA | NA | NA |
| 134 | Controls | Male | 21-30 | C | NA | NA | NA | NA | NA |
| 135 | Controls | Female | 31-40 | C | NA | NA | NA | NA | NA |
| 136 | Controls | Male | 41-50 | C | NA | NA | NA | NA | NA |
| 137 | Controls | Male | 21-30 | C | NA | NA | NA | NA | NA |
| 138 | Controls | Male | 21-30 | C | NA | NA | NA | NA | NA |
| 139 | Controls | Female | 21-30 | C | NA | NA | NA | NA | NA |
| 140 | Controls | Male | 21-30 | C | NA | NA | NA | NA | NA |
| 141 | Controls | Female | 21-30 | C | NA | NA | NA | NA | NA |
| 142 | Controls | Male | 21-30 | C | NA | NA | NA | NA | NA |
| 143 | Controls | Female | 51-60 | C | NA | NA | NA | NA | NA |
| 144 | Controls | Male | 51-60 | C | NA | NA | NA | NA | NA |
| 145 | Controls | Female | 21-30 | C | NA | NA | NA | NA | NA |
| 146 | Controls | Female | 31-40 | C | NA | NA | NA | NA | NA |
| 147 | Controls | Female | 31-40 | C | NA | NA | NA | NA | NA |
| 148 | Controls | Male | 31-40 | C | NA | NA | NA | NA | NA |
| 149 | Controls | Female | 21-30 | C | NA | NA | NA | NA | NA |
| 150 | Controls | Female | 51-60 | C | NA | NA | NA | NA | NA |
| 151 | Controls | Female | 21-30 | C | NA | NA | NA | NA | NA |
| 152 | Controls | Female | 21-30 | C | NA | NA | NA | NA | NA |
| 153 | Controls | Female | 21-30 | C | NA | NA | NA | NA | NA |
| 154 | Controls | Male | 41-50 | C | NA | NA | NA | NA | NA |
| 155 | Controls | Male | 31-40 | C | NA | NA | NA | NA | NA |
| 156 | Controls | Female | 21-30 | C | NA | NA | NA | NA | NA |
| 157 | Controls | Female | 21-30 | C | NA | NA | NA | NA | NA |
| 158 | Controls | Female | 21-30 | C | NA | NA | NA | NA | NA |
| 159 | Controls | Female | 21-30 | C | NA | NA | NA | NA | NA |
| 160 | Controls | Male | 31-40 | C | NA | NA | NA | NA | NA |
| 161 | Controls | Male | 31-40 | C | NA | NA | NA | NA | NA |
| 162 | Controls | Female | 21-30 | C | NA | NA | NA | NA | NA |
| 163 | Controls | Male | 31-40 | C | NA | NA | NA | NA | NA |
| 164 | Controls | Female | 21-30 | C | NA | NA | NA | NA | NA |
| 165 | Controls | Female | 31-40 | C | NA | NA | NA | NA | NA |

**Supplementary Table 2:** Domain Specific Neuropsychological Measures

| **Domain** | **List of Measures** |
| --- | --- |
| **Language** | 1. RBANS: Picture Naming 2. Boston Naming Test 3. NAB Naming Test 4. Semantic/Category Fluency 5. Phonemic/Letter Fluency 6. RBANS: Semantic Fluency 7. D-KEFS: Letter Fluency 8. D-KEFS: Category Fluency |
| **Memory** | 1. RBANS: List Recall 2. RBANS: Story Recall 3. RBANS: Figure Recall 4. WMS-IV Logical Memory II 5. NAB Story Learning Phrase Unit Delayed Recall 6. WMS-IV Visual Reproduction II 7. WMS-IV or WMS-III Faces II 8. California Verbal Learning Test-2^nd^ or 3^rd^ editions (CVLT) Scores: delayed free recall 9. Hopkins Verbal Learning Test-Revised (HVLT-R) Scores: delayed recall 10. Rey-Osterrieth Complex Figure (Delayed Recall) 11. Recognition Memory Test (RMT) Words and Faces 12. Brief Visual Memory Test-R (BVMT-R) Delayed Recall |
| **Executive Function** | 1. Trail Making Test B (visual or oral version) 2. D-KEFS: Number-Letter Sequencing 3. WAIS-IV or WAIS-5 or WASI-II- Matrix Reasoning 4. WAIS-IV or WAIS-5 or WASI-II - Similarities 5. WAIS-IV or WAIS-5- Figure Weights 6. 20 Questions (D-KEFS subtest) |
| **Visual Spatial Perception** | 1. Block Design (WAIS-IV or WAIS-5 or WASI-II subtest 2. Judgment of Line Orientation (JOLO) 3. RBANS: Line Orientation 4. Benton Facial Recognition Test 5. Rey Complex Figure (copy) 6. RBANS: Figure Copy 7. Visual Puzzles Subtest (WAIS-IV or WAIS-5) |

**Supplementary Table 2:** Association between Hub disruption indices with clinical variables (Values indicate FDR corrected *p* value)

| **Clinical variable** | ***K_s_*** | ***K_BC_*** | ***K_CC_*** | ***K_GE_*** |
| --- | --- | --- | --- | --- |
| Age at scan (yrs) | 0.989 | 0.985 | 0.985 | 0.985 |
| Sex, male (%) | 0.985 | 0.985 | 0.989 | 0.989 |
| MRI lesional (%) | 0.989 | 0.985 | 0.989 | 0.989 |
| Lateralization | 0.989 | 0.989 | 0.985 | 0.985 |
| Seizure location | 0.989 | 0.985 | 0.989 | 0.989 |
| Age of onset (yrs) | 0.989 | 0.989 | 0.989 | 0.995 |
| Disease duration (yrs) | 0.989 | 0.989 | 0.989 | 0.989 |
| Hippocampal sclerosis (%) | 0.989 | 0.989 | 0.989 | 0.989 |

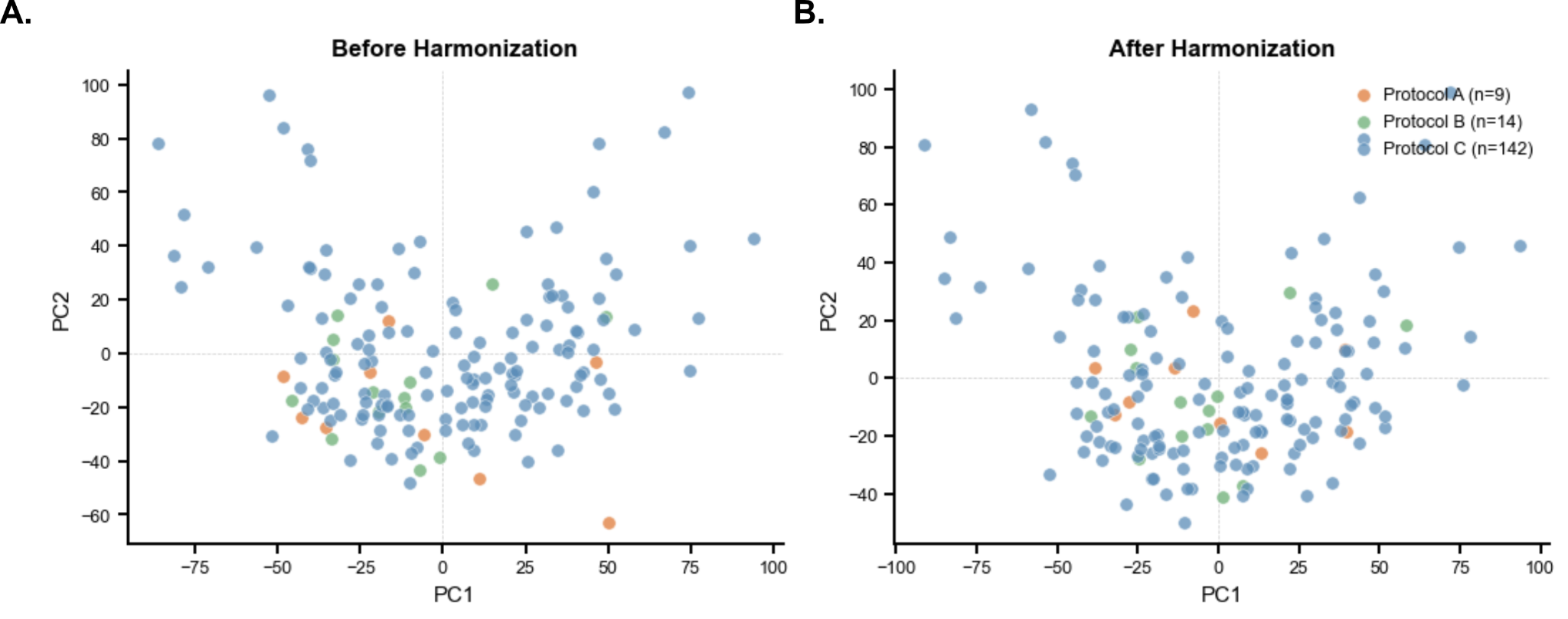

**Supplementary Figure S1: Effects of harmonization on functional connectivity in controls and epilepsy subjects:** Functional connectivity plotted in principal component space for each subject from protocol A, B and C, **A.** before and **B.** after harmonizing the functional connectivity matrix.

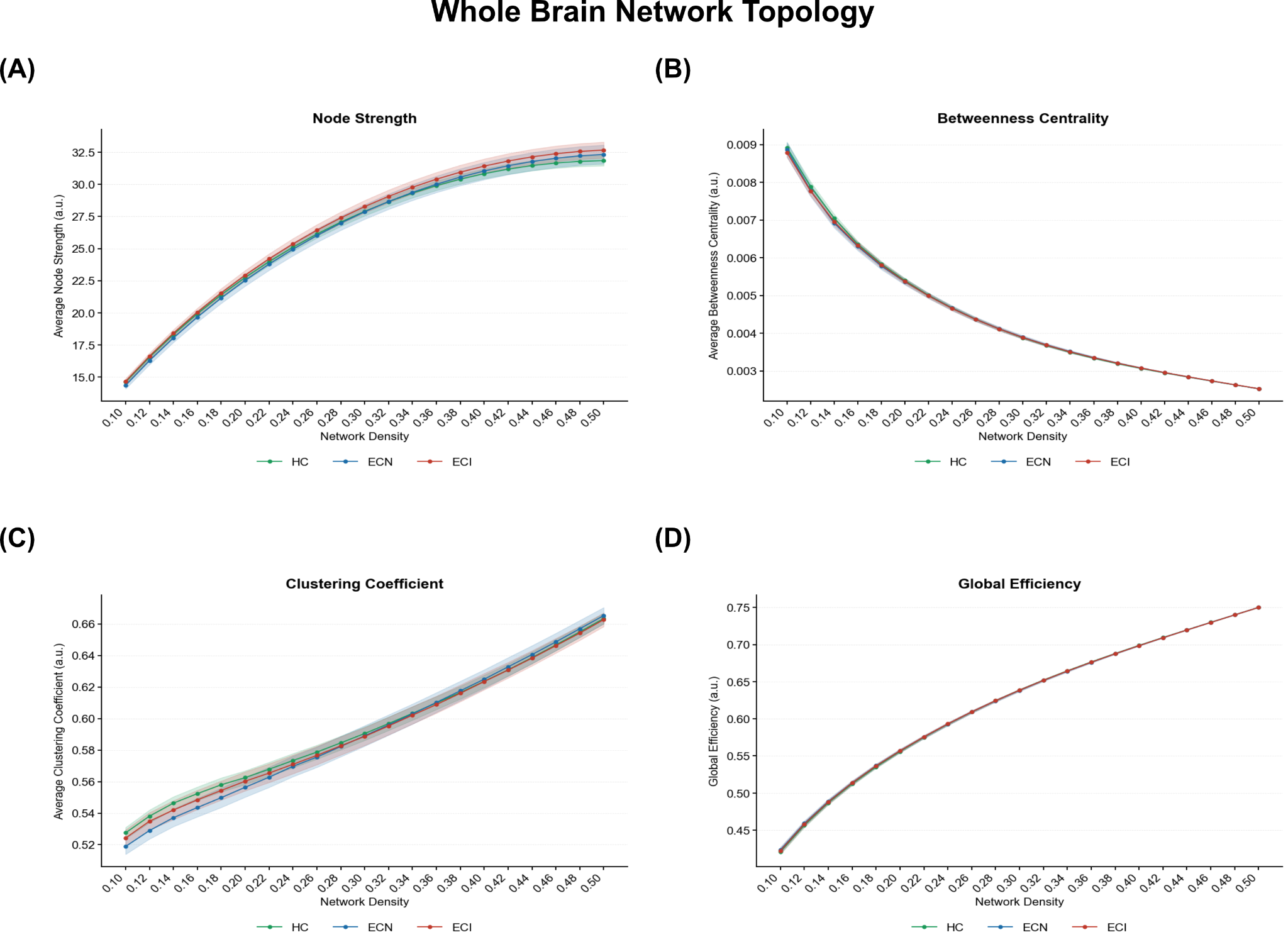

**Supplementary Figure S2:** Network metrics: Whole-brain **A.** average node strength, **B.** average betweenness, **C.** average clustering coefficient, and **D.** global efficiency from network densities 0.1 to 0.5 for HC (green), ECN (blue) and ECI (red). bold lines represent the mean; shaded curve represents 95% confidence interval (CI). HC: Healthy controls, ECN: Epilepsy with cognitively neutral, ECI: Epilepsy with cognitively impaired.

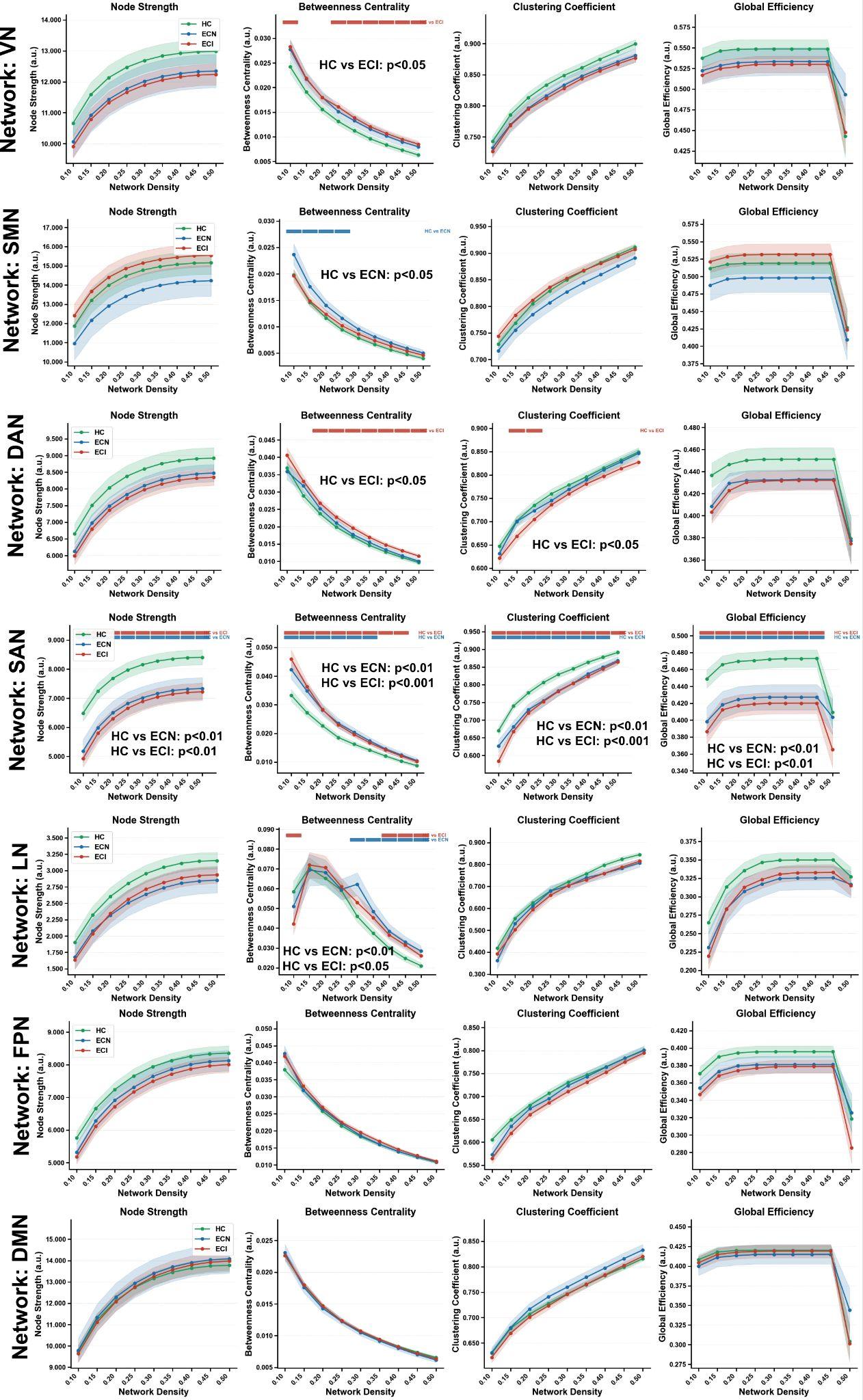

**Supplementary Figure S3: Intra-network topological properties across cognitive phenotypes.** Node strength , betweenness centrality, clustering coefficient, and nodal efficiency are shown for seven canonical resting-state networks at a connection densities ranging from 10-50% across HC, ECN, and ECI groups.

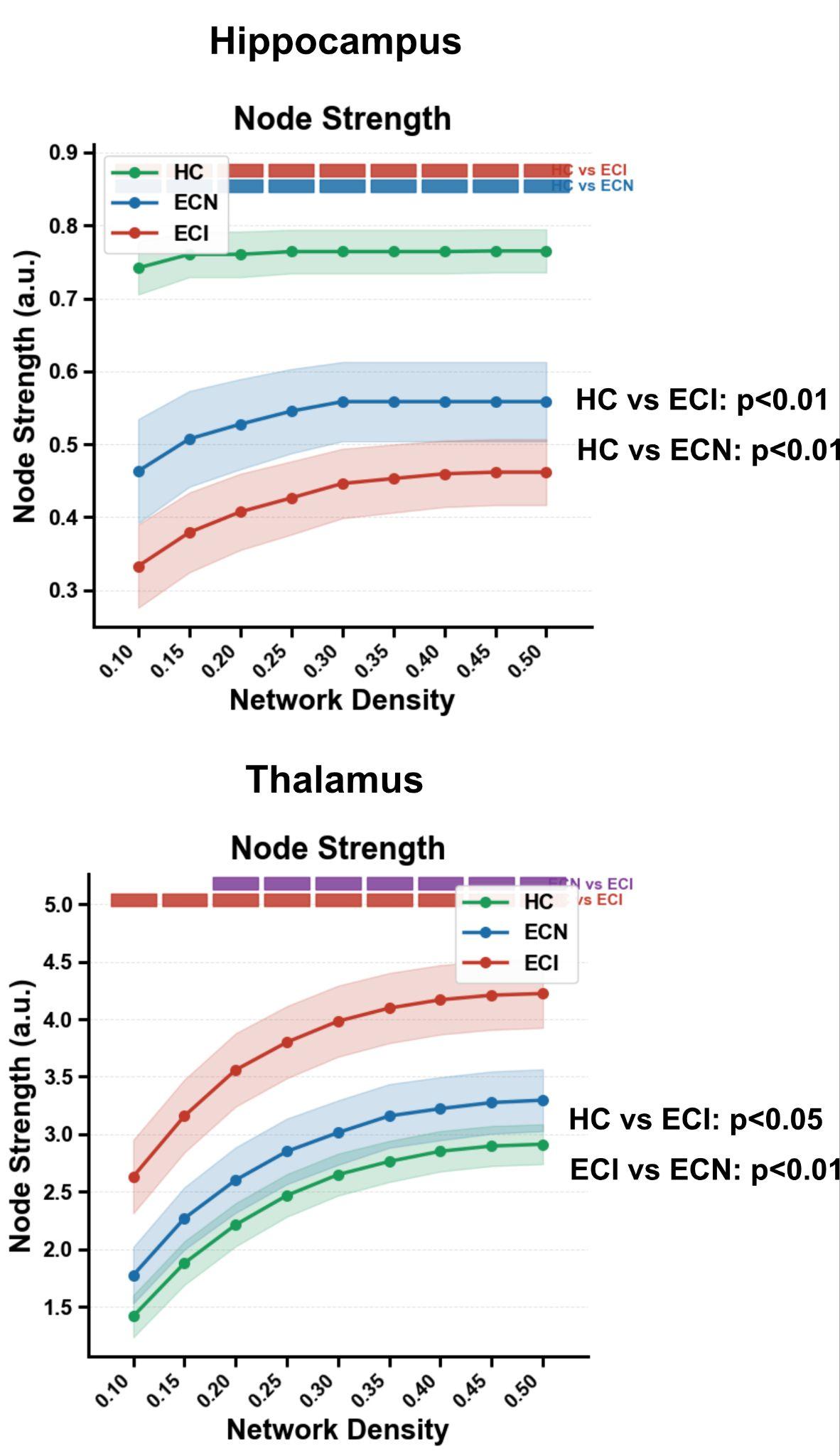

**Supplementary Figure S4. Inter-network functional connectivites across cognitive phenotypes.** Functional connectivities are shown for seven canonical resting-state networks at a connection density of 10% across HC, ECN, and ECI groups. Significant connectivity differences are shown using red and green chords in the chord diagram.

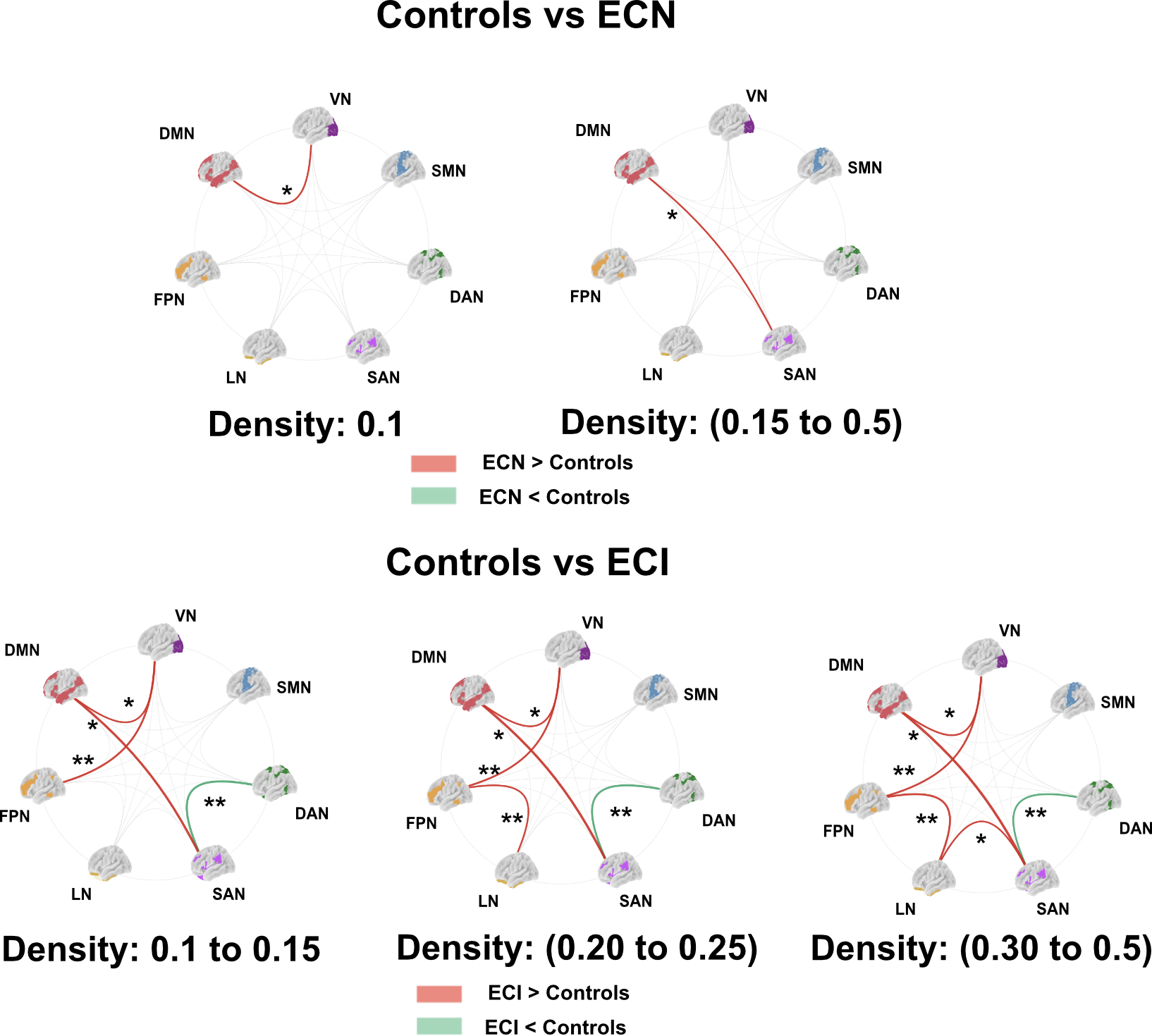

**Supplementary Figure S5. Inter-network functional connectivities across cognitive phenotypes.** Functional connectivities are shown for seven canonical resting-state networks at a connection density of 10% across HC, ECN, and ECI groups. Significant connectivity differences are shown using red and green chords in the chord diagram.

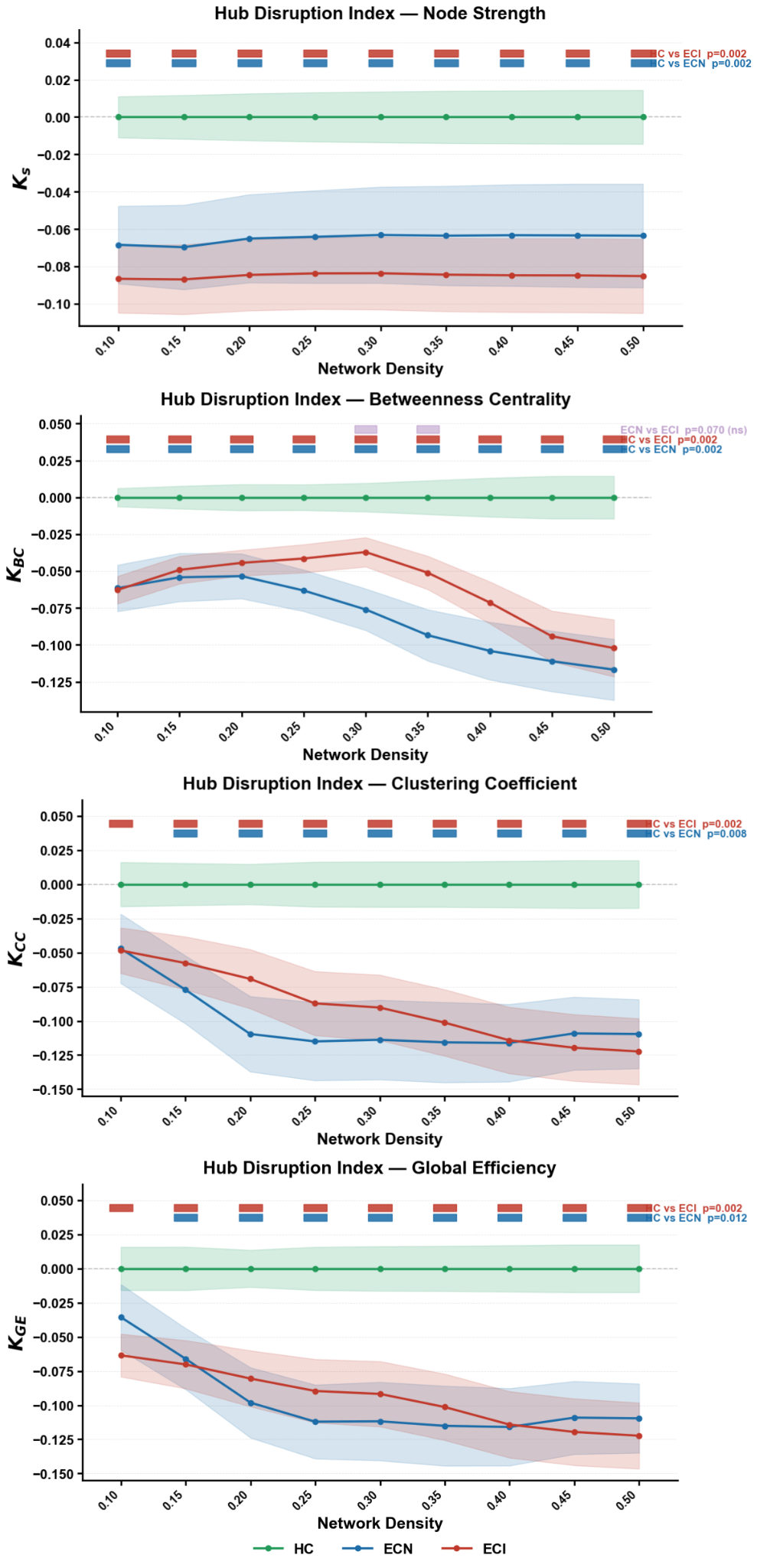

**Supplementary Figure S6: Hub disruption indices are stable across network density thresholds.** Sensitivity analysis of the hub disruption index computed across a range of graph densities (0.10–0.50) for four metrics. In each panel, lines show the group mean *K* as a function of network density for HC (HC, green), ECN (blue), and ECI (red), with shaded bands denoting the confidence interval. Colored bars at the top of each panel indicate the densities at which a pairwise comparison was significant, with *p*-values.

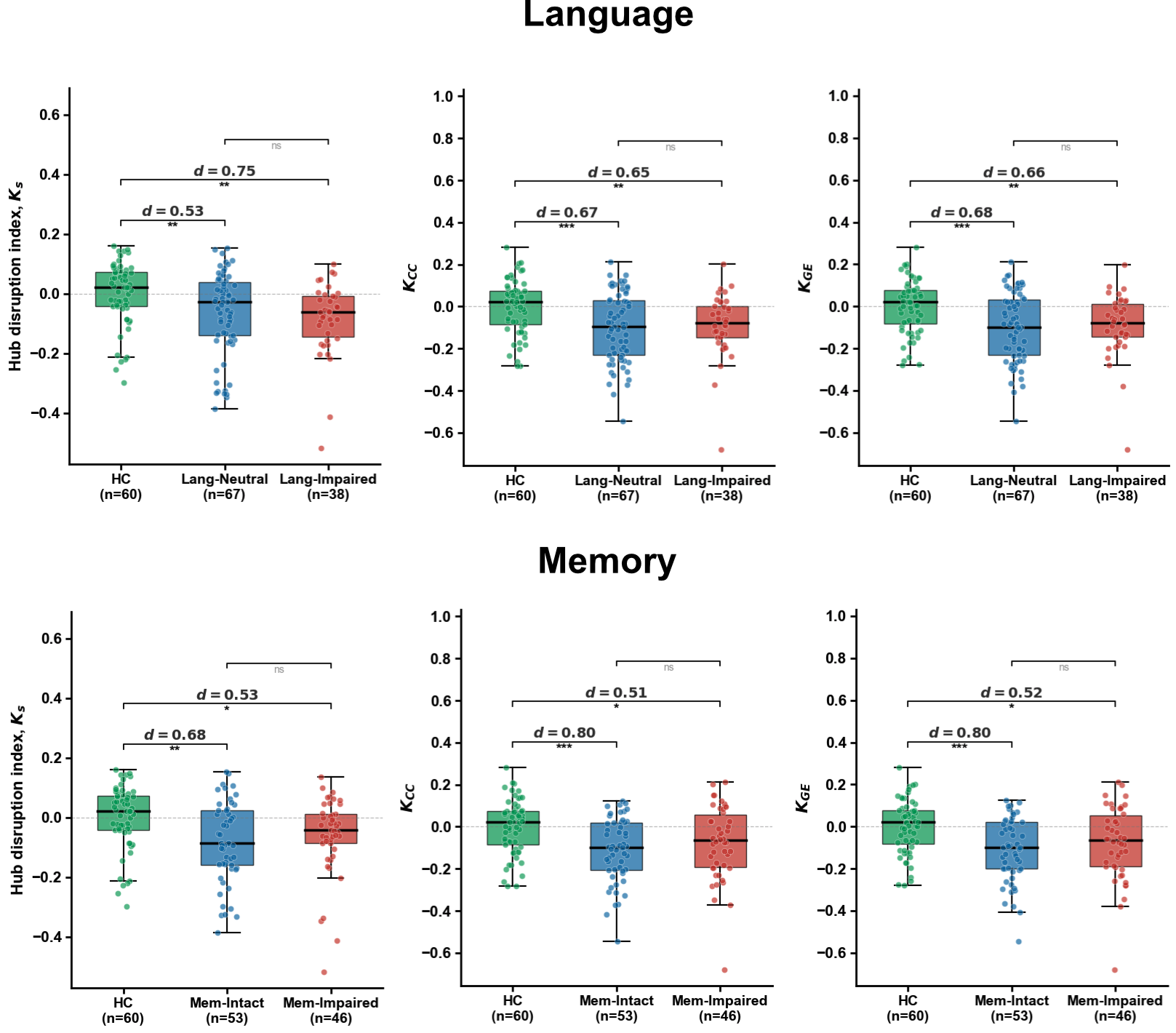

**Supplemental Figure S7. Domain-specific hub disruption indices for node strength, clustering coefficient, and global efficiency** **for the language (top) and memory (bottom) phenotypes** (language: HC *n* = 60, Lang-Neutral *n* = 67, Lang-Impaired *n* = 38; memory: HC *n* = 60, Mem-Intact *n* = 53, Mem-Impaired *n* = 46). Pairwise Cohen's *d* values are shown above each comparison. Significance: ns, not significant; **p* < 0.05, ***p* < 0.01, ****p* < 0.001.

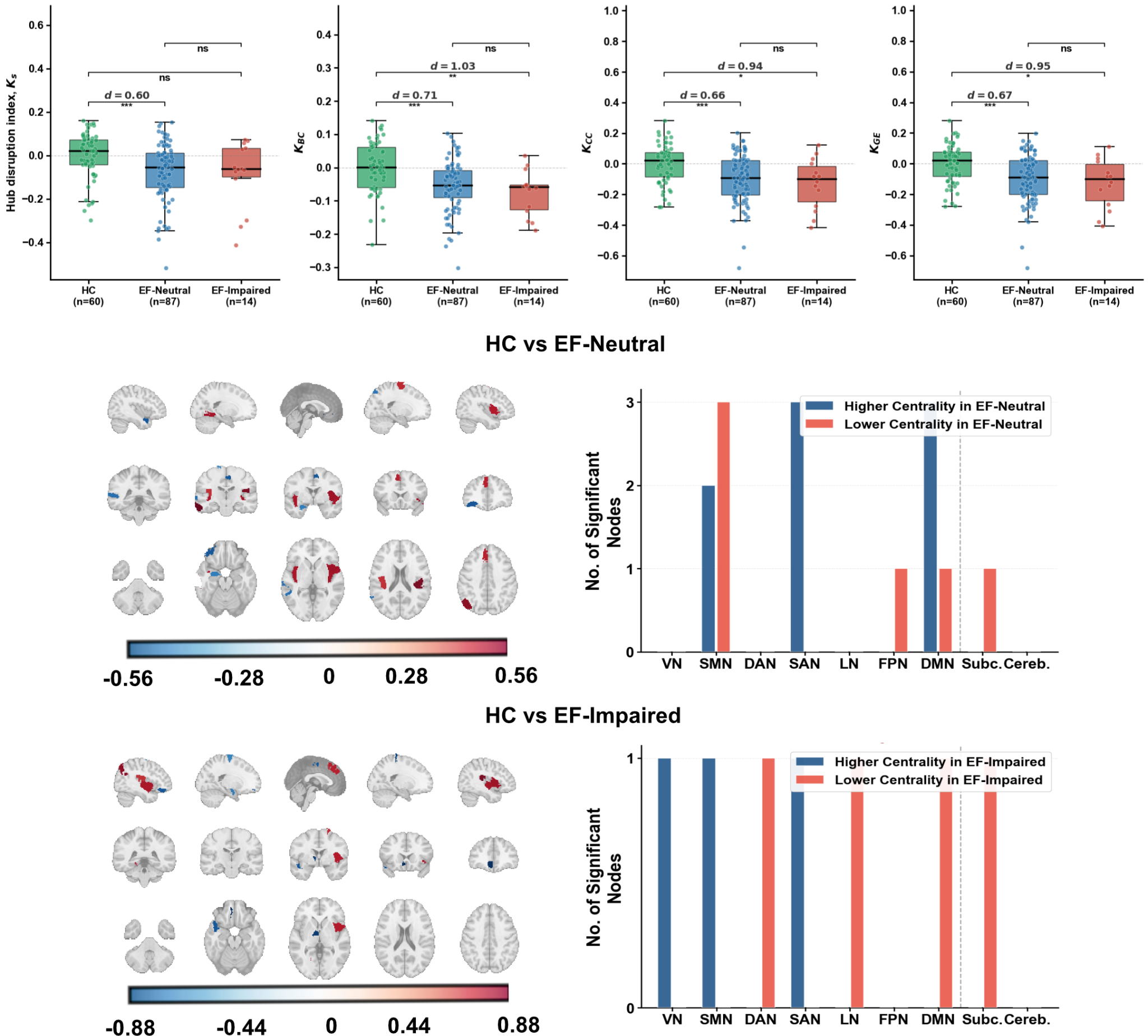

**Supplemental Figure S8. Executive-function (EF) phenotype: hub disruption indices and betweenness-centrality reorganization. (Top)** Group distributions of the four hub disruption indices ; node strength (*K*_S_), betweenness centrality (*K*_BC_), clustering coefficient (*K*_CC_), and global efficiency (*K*_GE_) for healthy controls (HC, *n* = 60) and patients split by executive-function performance into neutral (EF-Neutral, *n* = 87) and impaired (EF-Impaired, *n* = 14) subgroups. **(Middle and bottom)** Node-level betweenness-centrality reorganization relative to controls for HC vs. EF-Neutral (middle) and HC vs. EF-Impaired (bottom). Brain panels project parcels with significant betweenness-centrality differences onto sagittal, coronal, and axial MNI slices, colored by direction (blue, higher centrality in the patient subgroup; red, lower; per-comparison scales shown below each set). The bar charts count significant nodes per network — visual (VN), somatomotor (SMN), dorsal attention (DAN), salience (SAN), limbic (LN), frontoparietal (FPN), default mode (DMN), subcortical (Subc.), and cerebellar (Cereb.) — split by direction. Significance: ns, not significant; **p* < 0.05, ***p* < 0.01, ****p* < 0.001.

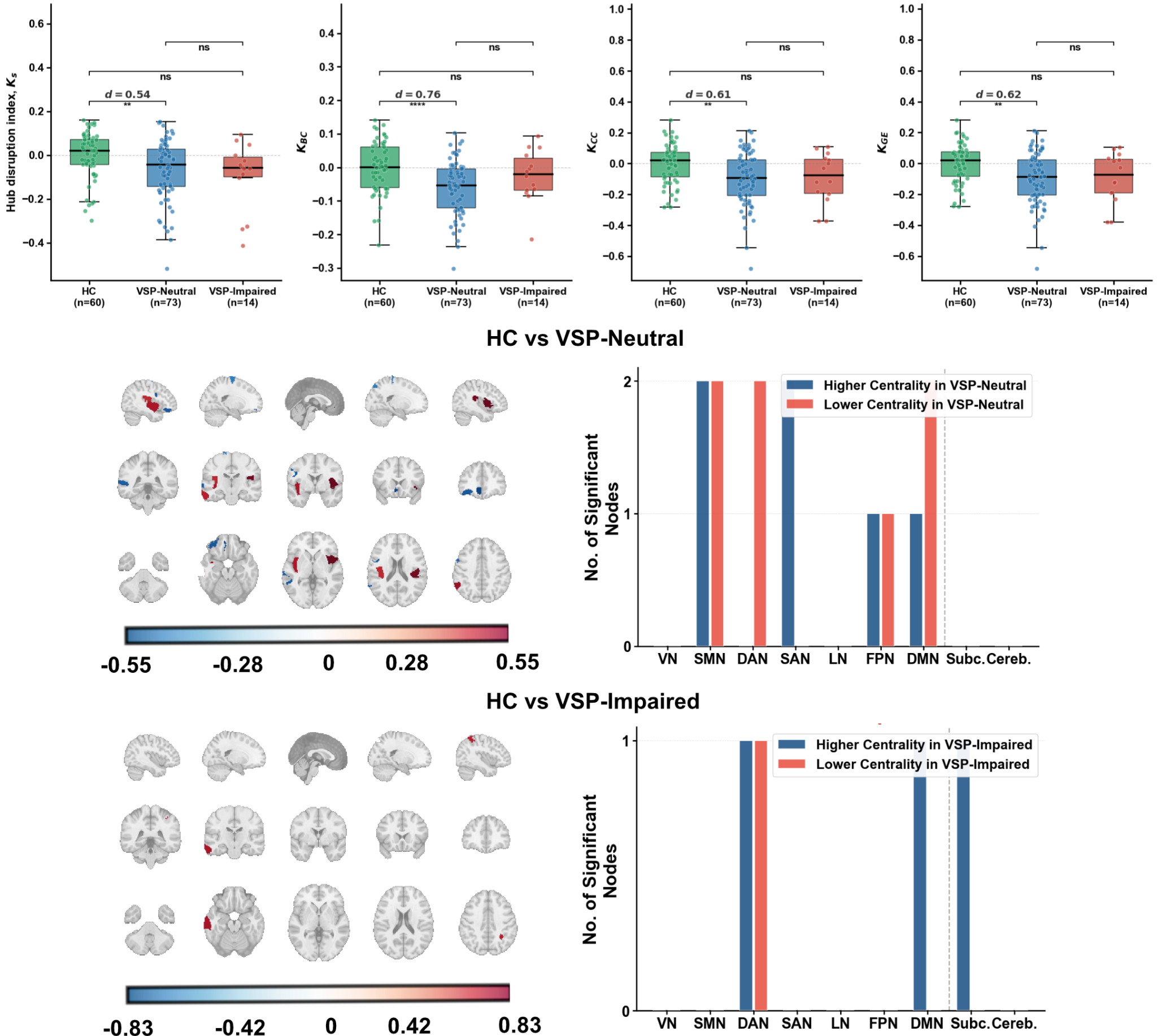

**Supplemental Figure S9. Visual Spatial Perception (VSP) phenotype: hub disruption indices and betweenness-centrality reorganization. (Top)** Group distributions of the four hub disruption indices ; node strength (*K*_S_), betweenness centrality (*K*_BC_), clustering coefficient (*K*_CC_), and global efficiency (*K*_GE_) for healthy controls (HC, *n* = 60) and patients split by VSP performance into neutral (VSP-Neutral, *n* = 87) and impaired (VSP-Impaired, *n* = 14) subgroups. **(Middle and bottom)** Node-level betweenness-centrality reorganization relative to controls for HC vs. VSP-Neutral (middle) and HC vs. VSP-Impaired (bottom). Brain panels project parcels with significant betweenness-centrality differences onto sagittal, coronal, and axial MNI slices, colored by direction (blue, higher centrality in the patient subgroup; red, lower; per-comparison scales shown below each set). The bar charts count significant nodes per network — visual (VN), somatomotor (SMN), dorsal attention (DAN), salience (SAN), limbic (LN), frontoparietal (FPN), default mode (DMN), subcortical (Subc.), and cerebellar (Cereb.) — split by direction. Significance: ns, not significant; **p* < 0.05, ***p* < 0.01, ****p* < 0.001.
